# Essential hypertension is associated with changes in gut microbial metabolic pathways: A multi-site analysis of ambulatory blood pressure

**DOI:** 10.1101/2021.02.18.21252018

**Authors:** Michael Nakai, Rosilene V Ribeiro, Bruce R. Stevens, Paul Gill, Rikeish R. Muralitharan, Stephanie Yiallourou, Jane Muir, Melinda Carrington, Geoffrey A. Head, David M. Kaye, Francine Z. Marques

**Affiliations:** Hypertension Research Laboratory, School of Biological Sciences, Monash University, Melbourne, Australia; Charles Perkins Centre, University of Sydney, Sydney, Australia; Department of Physiology, Monash University, Melbourne, Australia; School of Life and Environmental Sciences, Faculty of Science, The University of Sydney, Sydney, Australia; University of Florida, College of Medicine, Department of Physiology and Functional Genomics, Gainesville, FL, USA; Department of Gastroenterology, Monash University, Melbourne, Australia; Institute for Medical Research, Ministry of Health Malaysia, Kuala Lumpur, Malaysia; Preclinical Disease and Prevention, Baker Heart and Diabetes Institute, Melbourne, Australia; Neuropharmacology Laboratory, Baker Heart and Diabetes Institute, Melbourne, Australia; Department of Pharmacology, Faculty of Medicine Nursing and Health Sciences, Monash University, Melbourne, Australia; Heart Failure Research Group, Baker Heart and Diabetes Institute, Melbourne, Australia; Department of Cardiology, Alfred Hospital, Melbourne, Australia; Central Clinical School, Faculty of Medicine Nursing and Health Sciences, Monash University, Melbourne, Australia

**Keywords:** Hypertension, ambulatory blood pressure, gut microbiota, gut microbiome, gut metabolites, metabolites, short-chain fatty acids

## Abstract

Recent evidence supports a role for the gut microbiota in hypertension, but whether ambulatory blood pressure (BP) is associated with gut microbiota and their metabolites remains unclear. We characterised the function of the gut microbiota, their metabolites and receptors in untreated human hypertensive participants in Australian metropolitan and regional areas. Ambulatory BP, faecal microbiome predicted from 16S rRNA gene sequencing, plasma and faecal metabolites called short-chain fatty acid (SCFAs), and expression of their receptors were analysed in 70 untreated and otherwise healthy participants from metropolitan and regional communities. Most normotensives were female (66%) compared to hypertensives (35%, P<0.01), but there was no difference in age between the groups (59.2±7.7 versus 60.3±6.6-years-old). Based on machine-learning multivariate covariance analyses of de-noised amplicon sequence variant prevalence data, we determined that there were no significant differences in predicted gut microbiome α- and β-diversity metrics between normotensives versus essential, white coat or masked hypertensives. However, select taxa were specific to these groups, notably *Acidaminococcus spp., Eubacterium fissicatena and Muribaculaceae* were higher, while *Ruminococcus* and *Eubacterium eligens* were lower in hypertensives. Importantly, normotensive and essential hypertensive cohorts could be differentiated based on gut microbiome gene pathways and metabolites. Specifically, hypertensive participants exhibited higher plasma acetate and butyrate, but their immune cells expressed reduced levels of SCFA-activated G-protein coupled re^1^ceptor 43 (GPR43). In conclusion, gut microbial diversity did not change in essential hypertension, but we observed a significant shift in microbial gene pathways. Hypertensive subjects had lower levels of GPR43, putatively blunting their response to BP-lowering metabolites.

## Introduction

Lifestyle and environmental factors are well-established contributors to increased blood pressure (BP). The gut microbiota, the community of microorganisms that inhabit our small and large intestines, is a newly described risk factor for the development of hypertension.^2, 3^ Importantly, increasing evidence using faecal microbiota transplants (FMTs) to germ-free animals supports that the gut microbiota is not merely associated with experimental and essential hypertension, but it drives an increase in BP.^4–6^ While some studies have explored the association between essential hypertension and the gut microbiota,^5, 7–14^ a major limitation has been the use of self-reported hypertension or office BP, which do not allow for distinction between types of hypertension and decrease the power of studies.^2^ All but one study also included patients using a varied range of BP-lowering medications, but each of these medications on their own have been reported to affect the gut microbiota.^15, 16^

A major modulator of the gut microbiota is diet, which can alter the microbiota in as little as a few days.^17^ Dietary fibre, in particular soluble fibre and resistant starches (also known as prebiotics), are particularly important as they feed commensal bacteria. Fibre is also important for BP control: in a meta-analysis and a prospective study that involved 388,000 participants followed-up for approximately 9 years, fibre intake was associated with lower risk of both cardiovascular and all-cause death, and lower systolic BP (SBP).^18^ This was validated by a more recent study, which identified that fibre intake is an underlying factor for mortality, driven by BP.^19^ Fermentation of fibre by gut bacteria results in the release of short-chain fatty acids (SCFAs). SCFAs, such as acetate, propionate and butyrate, are able to lower BP in experimental models of hypertension.^4, 9, 20, 21^ Yet, a lack of consistency in SCFA levels still exists across studies in essential hypertension and its association with BP.^12, 26^ This may be due to the volatile characteristic of these metabolites, making them challenging to quantify, as well as differences in colonic (i.e., produced), faecal (i.e., excreted) and circulatory (i.e., absorbed) levels.^27^

Here we aimed to characterise the predicted gut bacteriome, their function and levels of SCFAs in participants will well-characterised BP. Our study addresses the following limitations in our field: it is the first multi-site gut microbiota study which analysed samples from both male and female participants from regional and metropolitan areas, diagnosed using ambulatory BP monitoring (ABPM). As such, we provide the predicted gut bacteriome of patients with essential, masked and white coat hypertension. Importantly, all participants included were not receiving any BP-lowering medication. We also determined the impact of their diet on the predicted gut bacteriome and associated circulating and faecal levels of SCFAs and SCFA-sensing receptors.

## Methods

### Participants and recruitment

In this observational, case-control study, 76 participants were recruited from the community in metropolitan (n=41 Baker Institute and Alfred Hospital, Melbourne) and regional (n=35, Shepparton) areas (Figures S1 and S2) between October-2016 and April-2018. Inclusion criteria were: aged 40-70 years, either sex, body mass index (BMI) 18.5-30 kg/m^2^, and not using BP-lowering medication. Exclusion criteria included gastrointestinal disease (including history of intestinal surgery, inflammatory bowel disease, celiac disease, lactose intolerance, chronic pancreatitis or other malabsorption disorder), diabetes (type 1 and 2), chronic kidney disease, probiotics or antibiotics use in the past 3 months. Two participants were excluded due to high BMI and four were excluded due to incomplete 24-hour BP measurements. After exclusions, a total of 70 participants remained: 40 in the metropolitan clinics and 30 in the regional clinic (Figures S1 and S2). This study complied with the Declaration of Helsinki, and was approved by the human research ethics committee of the Alfred Hospital (approval 415/16). All participants provided informed consent. The study is registered in the Australian New Zealand Clinical Trials Registry under ACTRN12620000958987.

### Blood pressure measurement and hypertension diagnosis

Office BP was measured using an automatic BP monitor (Omron) after the participant had been seated for at least 5 minutes with both feet flat on the floor, legs uncrossed and upper arm relaxed. Two measurements were taken 1 minute apart, and if systolic BP was >15 mmHg apart of diastolic BP >10 mmHg apart a third measurement was performed and averaged. Participants were then fitted with a calibrated ABPM device (AND or SpaceLabs) for 24-hours and given clear instructions of how to operate the device and when to remove it. Hypertension was diagnosed using the European guidelines.^22^

### Food frequency questionnaire

Dietary intake over a period of 12 months was assessed using the Dietary Questionnaire for Epidemiological Studies (DQES) version 3.2, a self-administered and validated food frequency questionnaire (FFQ), developed by the Cancer Council Victoria that reflects dietary intake of the Australian population.^23^ Dietary intake estimates of 98 nutrients were derived from two Australian databases, AUSNUT 2007^24^ and NUTTAB 2010.^25^ Since data on fibre subtypes (insoluble fibre, soluble fibre and resistant starch) are not provided by DQES v3.1, we have followed the protocol previously described^26^ to obtain those estimates. This method involves calculating the amount of total fibre and fibre subtypes for each single food using the following sources: (1) latest dietary fibre values found on the AUSNUT 2011-13 database;^27^ (2) food industry provided dietary fibre composition data published by FSANZ^28^ and (3) the published dietary fibre data by the FSANZ in “Composition of Foods”, Australia, 1989.^29^

Participants’ diet quality was measured by comparing their intake (according to age and sex) with the Dietary Guidelines for Australian (DGA)^30^ recommendations using the Australian Dietary Guideline Index (DGI-2013).^31, 32^ The DGI-2013 includes ten components in total: six core-food components reflecting adequacy, quality and variety of intake within the ADG core food groups (vegetables, fruit, grains, lean meats and alternatives, and dairy and alternatives), one reflecting adequacy of fluid intake, and three noncore food groups (unsaturated spreads or oils, discretionary items and alcohol) reflecting compliance with guidelines to moderate or limit intake. The quality of grain (prefer wholegrain), dairy foods (prefer reduced-fat) and fluid intake (prefer water) was also assessed to reflect the recommendations set by ADG. Total beverage intake consisted of water, fruit juice, tea, coffee, milk and milk-based beverages.^30, 33^ Altogether, a maximum of 120 points could be achieved if all ADG recommendations were met (i.e. high diet quality).

### Faecal DNA extraction, library preparation and sequencing

Our study followed guidelines for gut microbiota studies in hypertension,^34^ and the Strengthening The Organization and Reporting of Microbiome Studies (STORMS) reporting (Table S1, Figure S2).^35^ Stool samples were collected by participants at home in empty tubes (for SCFA determination) or tubes containing RNAlater (Thermo Scientific, previously shown to preserve bacterial DNA^36^) for microbial DNA extraction. Tubes were brought to the clinics immediately or stored at -20°C for less than 24-hours and then brought to the clinics, where they were stored at -80°C until further processing. DNA was extracted using the DNeasy PowerSoil DNA isolation kit (Qiagen). The V4-V5 region of the bacterial 16S rRNA was amplified by PCR using 20ng of DNA, Platinum Hot Start PCR master mix (ThermoFisher Scientific), 515F and 926R primers (Bioneer), single-indexing and methods previously described in the Earth Microbiome Project^37^ in a Veriti Thermal Cycler (ThermoFisher Scientific). As such, all the gut microbiome data reported refers to predicted faecal microbiome, as it was not obtained from shot-gun metagenome sequencing. Non-template controls (NTC) were used to identify contamination. The quality and quantity of the PCR product was assessed in a Qubit (ThermoFisher Scientific). No NTC samples showed any amplification. 240ng of PCR product per sample were pooled and cleaned using the PureLink PCR Purification kit (ThermoFisher Scientific). The product was then sequenced in a Illumina MiSeq sequencer (300 bp paired-end reads) with 20% PhiX spiked in. To increase the reproducibility of the findings, all samples were independently sequenced twice. These technical duplicated samples were combined for the analyses described below.

### Bioinformatic analyses of gut microbiome

Sequence reads from samples were first analysed using the QIIME2 framework.^38^ Forward and reverse reads were first truncated at base number 243 for forward reads and base number 224 for reverse reads, then were denoised, merged, and chimera filtered using the DADA2 plugin,^39^ resulting in an amplicon sequence variant (ASV) table (via q2-dada2) with resolution at the single nucleotide level. A phylogeny was then created using fasttree2^40^ from mafft-aligned^41^ ASVs, and were then subsampled without replacement to 29,000 reads per sample (via q2-alignment and q2-phylogeny). Both α and β diversity metrics were generated from the rarefied samples (via q2-diversity), including unweighted and weighted Unifrac metrics along with associated Principle Coordinate Analysis (PCoA) distance tables. Taxonomic assignment used a nai□ve Bayes classifier to label ASVs (via q2-feature-classifier^42^), trained against the SILVA database (version 138) 99% OTU reference sequences specific for bacterial V4-V5 rRNA regions. Categorical sample metadata predictions were attempted on the rarefied samples using the q2-sample-classifier plugin.^43^ Linear discriminant analysis (LDA) effect size (LEfSe)^44^ was used to identify differentially abundant taxa between groups, with a specified effect size cut-off of 2.0 and Kruskal-Wallis test *P*<0.05. This data was then imported to Calypso^45^ and analysed using Spearman correlations to validate these associations with BP as a continuous variable using the top 200 most abundant taxa. These analyses were adjusted for multiple comparisons using false discovery rate (FDR). Microbiome data can be accessed at the NCBI Sequence Read Archive (SRA) database with assess numbers SAMN18752981 through SAMN18753049.

Separately, the OTU table output from the QIIME2 framework was converted to predicted gene abundances on the Piphillin^46^ and PICRUSt2 servers. These servers do not cover the same pathways, thus some specific pathways are not represented in both and they complement each other. The resulting hypothesised gene abundances were then imported into R (version 4.0.2) using the package phyloseq^47^ and transferred through the R package PIME^48^ to capture gene abundance differences only at high sample prevalence levels. Effect sizes for genes were generated when PIME-filtered tables were passed onto ALDEx2, which conducted a differential abundance analysis comparing samples grouped by selected categorical variables in the metadata. ALDEx2 also inferred biological and sampling variation to calculate the expected false discovery rate, based on a Welch’s t-test.^49^ Via the DADA2/PIME/ALDEx2 pipeline, we used denoised prevalences which permit valid permutational multivariate analyses of covariance-based dissimilarities and effect sizes using a compositional dataset approach. To this end, in the present study we employed a machine learning algorithm using that produced denoised prevalences at the level of single nucleotide resolution. Our study benefited from our machine learning workflow that performed an internal validation of prevalence data that accounts for power, and reports significant differences—subtle or large—with respect to normalization, eigenvalue ordination, permutational analyses of covariance, dissimilarity matrices, and correlation with clustering within the 95% Confidence Interval, shown as principal coordinate analyses (PCA) plots, to show differences between groups.

### Short-chain fatty acids measurement

Plasma SCFAs were measured in 200 µL and faecal SCFAs were measured from 1 g of faecal sample, all in triplicates, as previously published.^50, 51^ Heptanoic acid was used as an internal standard. Samples were analysed using an Agilent GC6890 coupled to a flame-ionisation detector (FID), with helium used as the carrier gas. An Agilent FFAP column (30 m x 0.53 mM (internal diameter) x 1.00 μM (film thickness)) was installed for analysis. A splitless injection technique was used, with 0.2 μL of sample injected. A constant flow rate of 4.0 mL/min was used on the column. Upon injection, the oven was initially held at 90 °C for 1 minute, then raised to 190 °C at 20 °C/min and held for 3 minutes. A coefficient of variation of <10% within triplicate samples was used as a quality control measure.

### Blood expression of SCFA receptors and transporters

We have previously shown in animal models that all 3 main SCFA-sensing receptors GPR41 (*FFAR3)*, GPR43 (*FFAR2*) and GPR109A (*HCAR2*) have a role in cardiovascular dysfunction.^3^ These receptors are highly expressed in immune cells.^52^ To determine whether hypertensive patients had a deficiency in sensing these receptors, we quantified their levels in white blood cells from the participants with the highest (n=22 hypertensives) and lowest BP (n=25). Blood RNA was extracted using the Red Blood Cell Lysis Buffer (ThermoFisher Scientific) and RNeasy Mini kit (Qiagen). RNA was quantified in a Nanodrop Spectrophotometer and first-strand complementary synthesis reaction (cDNA) was made using the SuperScript™ IV VILO™ Master Mix with ezDNase™ Enzyme (ThermoFisher Scientific). TaqMan assays (Table S2) and the TaqMan Advanced Fast Master Mix were used in a QuantStudio 6 Flex Real-Time PCR (qPCR) system (all ThermoFisher Scientific). Glyceraldehyde 3-phosphate dehydrogenase (*GAPDH*) and β-actin (*ACTB*) were used as housekeeping genes. All expression experiments were run in duplicates and significance was assessed by 2^-ΔΔ*C*T^method.

### Statistical analyses

Microbiome data was analysed as explained above. Data were analysed blind. GraphPad Prism (version 8) package was used for statistical analysis and graphing. Results were tested for outliers using the Robust regression and outlier removal (ROUT) method^53^ and for normal distribution using the D’Agostino & Pearson tests. Non-parametric tests were used in case of non-normally distributed data. An *F* test was used to verify similar variance between groups. Two-tail independent sample t-tests (with Welch’s correction in the case of different variance) were used to compare demographic, BP, qPCR and SCFA data between 2 groups. Correlations were performed using Pearson’s or Spearman’s correlation coefficients. Further analyses were conducted using step-wise multiple linear regression models for acetate, butyrate and *GPR43* levels. These models had clinical (age, sex, body mass index, recruitment centre) variables as independent parameters (criteria of F-entry probability: 0.15, removal: 0.20) in Statistical package SPSS for Windows (release 25). Demographic, BP, qPCR and SCFA data are presented as mean±SEM, and those with a *P*<0.05 considered significant.

## Results

### Participants’ characteristics

A total of 70 participants were included in this study (Table 1). Based on ABPM, we identified that 33% of participants had hypertension. Four percent had white coat and 19% were masked hypertensives. Hypertensive patients had a similar age and body mass index (BMI) compared to normotensive controls, but there were sex differences between hypertensive, masked hypertensive and normotensive participants. As expected hypertensive and masked hypertensive participants also had higher ABPM, while white coat hypertensive participants had high office BP, but not ABPM. One participant was excluded due to low number of microbiome sequencing reads, resulting in a total of 69 participants included in the sequencing analyses below. Due to the small number of white coat hypertensives, we did not perform any predicted gut microbiome analyses in this group.

**Table 1.**
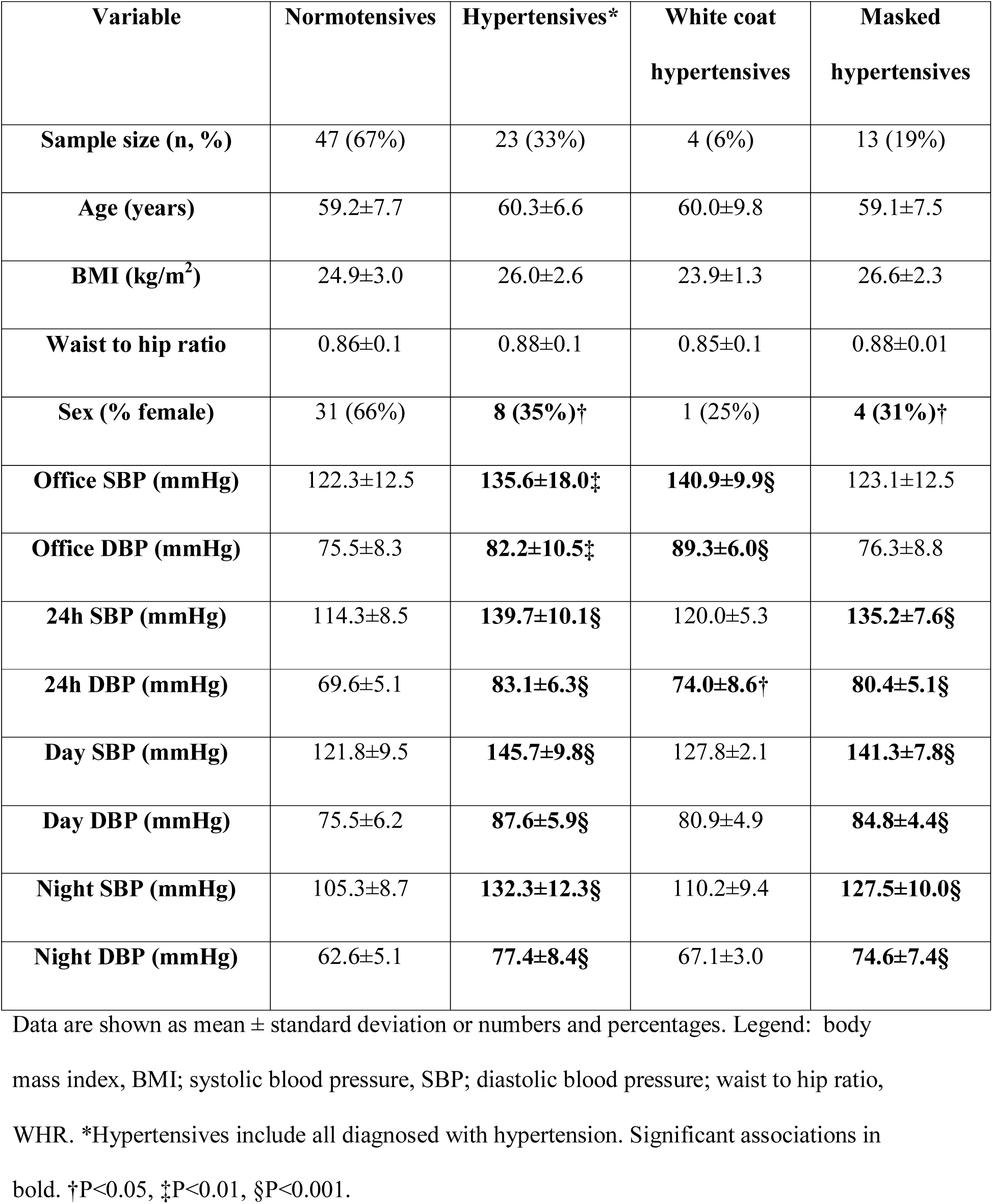
Demographics and clinical characteristics of participants.

### Dietary food intake

There was no difference between total fibre, resistant starches, insoluble and soluble fibre levels between normotensive and hypertensive participants, nor for different types of food individually that contribute towards the total fibre intake (e.g., nuts, legumes, grains) (Table S3). There was also no statistically significant difference in Australian dietary score between normotensive (60.2±1.1) and hypertensive participants (58.5±1.6, *P*=0.358).

### Gut microbiome

In total, over 4.3 million reads were denoised, merged, and survived chimera filtering, a survival rate of 43% with read counts averaging 63,000 reads per sample. Rarefaction of samples to 29,000 reads showed consistent and plateauing diversity metrics across samples (Figure S3), but caused the loss of data from one participant. We quantified two commonly used metrics for predicted gut microbiome studies, α- (i.e., a within sample metrics) and β-diversity (i.e., a between samples metrics). We found no association between 5 measurements of α-diversity and essential hypertension (Table S4) or masked hypertension in t-tests nor using machine learning algorithms (data not shown). We identified similar results when we studied β-diversity: there was no difference between normotensive controls and essential hypertensive (Figure S4) and masked hypertensive (Figure S5) participants. There was also no β-diversity difference between clinics, sex, age, BMI, fibre intake or diet score (Figure S6). However, some specific taxa were still more prevalent in hypertensive groups or normotensive controls, shown by LDA scores higher than 2 in normotensive or hypertensive groups (Figure 1 and Figure S7). This cut-off was chosen specifically to identify taxa that were likely to have a biological significance instead of just significant *P*-value. We identified that normotensive subjects had higher levels of *Ruminococcus spp.* and *Eubacterium eligens,* while essential hypertensive subjects had higher levels of *Acidaminococcus spp., Eubacterium fissicatena* and *Muribaculaceae spp* (Figure 1 and S7); masked hypertensives had higher levels of *Alistipes, Muribaculaceae,* (Figure 1). To validate these findings, we then performed correlations between the microbiome and 24-hour SBP and DBP (Figure S8, Table S5). *Acidaminococcus* was positively correlated with BP (r=0.28 to 0.34, *P*=0.005-0.018), and *Ruminococcus* (r=-0.26 to -0.32, *P*=0.008-0.031) and *Eubacterium eligens* (r=-0.28 to -0.3, *P*=0.013-0.021) negatively correlated with BP. However, no taxa reached FDR<0.05. Differently from previously reported,^54^ we did not identify any changes in *Lactobacillus spp*. in any of the groups nor associations with reported sodium intake (data not shown).

**Figure 1.**
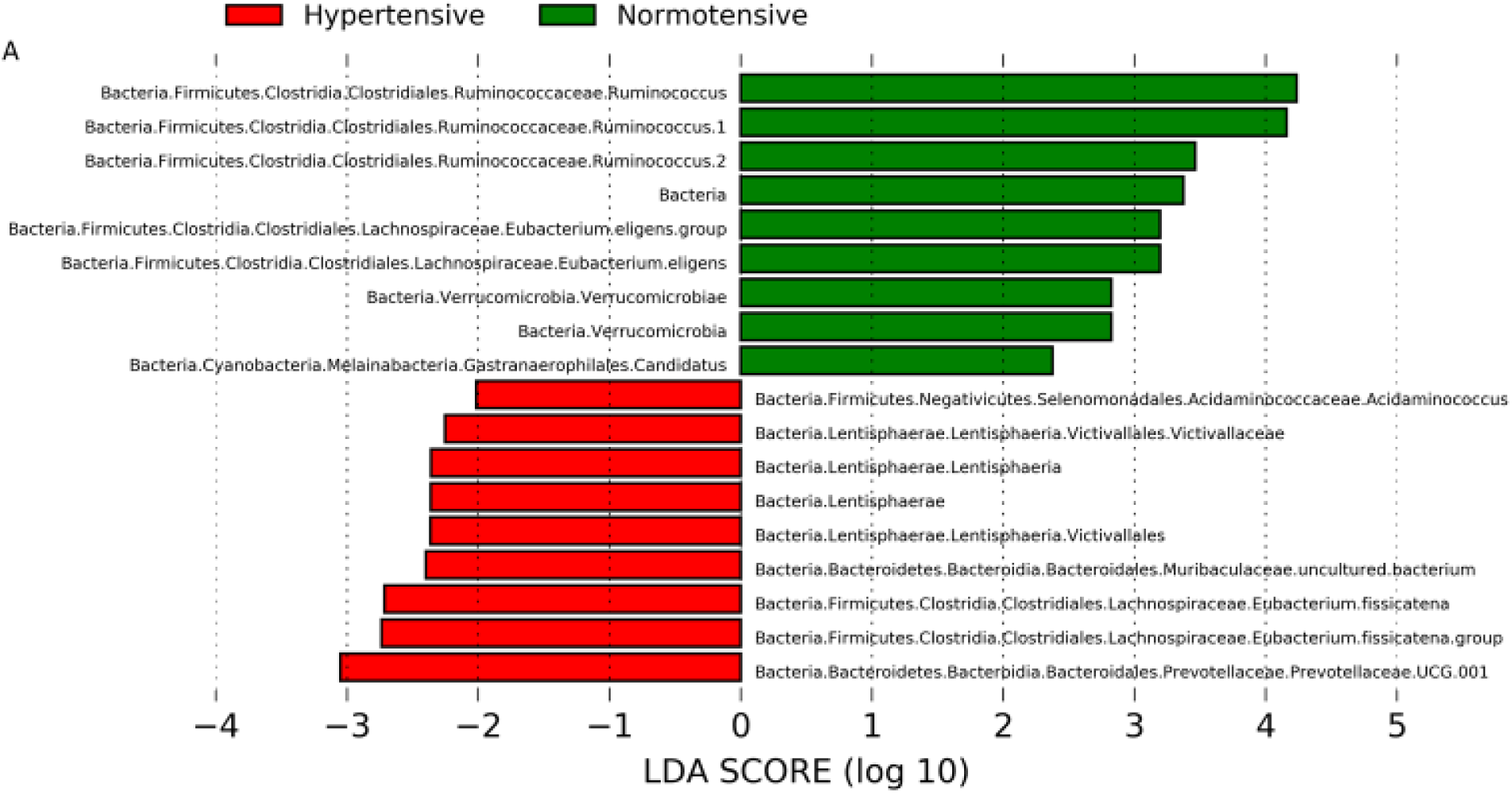
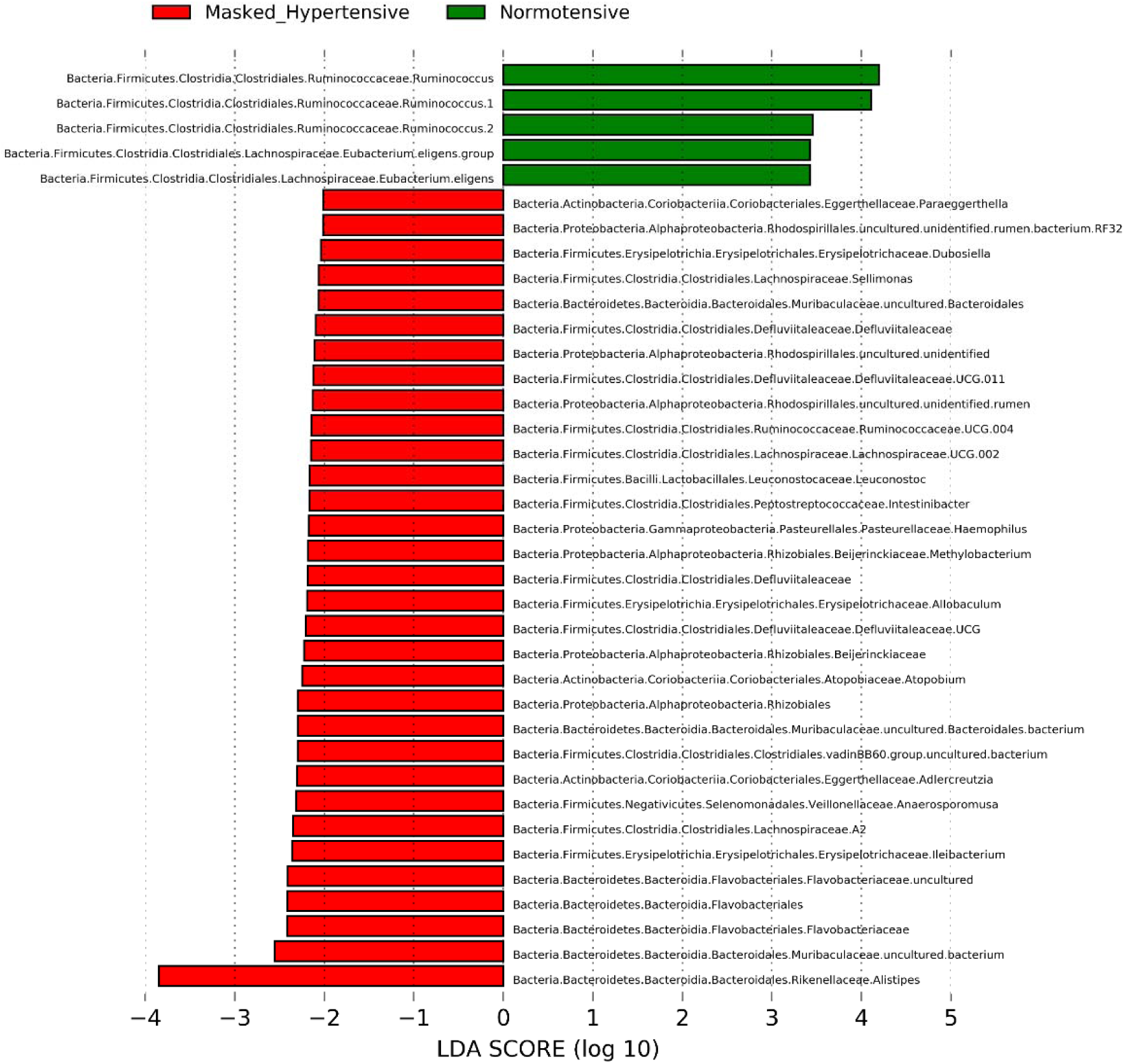
Predicted gut microbiome taxa linear discriminant analysis effect sizes. Predicted gut microbiome taxa that are different between normotensive subjects and A, essential hypertensives (n=46 normotensives and 23 hypertensives), B, masked hypertensives (n=46 normotensives and 13 masked hypertensives), with a linear discriminant analysis (LDA) score of least 2.

To determine whether predicted biological pathways with high sample prevalence rather than specific types of bacteria were associated with hypertension, we then analysed the data using ALDEx2 and PIME, which employs robust machine learning combined with microbiome pathway analysis. 4,038 predicted genes were identified after passing QIIME2-outputted ASV tables through the PICRUSt2 server (Figure 2) and 5,488 predicted genes in the Piphillin server (Figure S9). The following results are described for the PICRUSt2 server, although the Piphillin server generated similar results (Figure S9). After filtering out low sample prevalence genes using PIME (Figure 2A and 2B) and finding the prevalence interval with the lowest out-of-bag error rate (Figure 2C), only 2,339 genes and 3,950,619,227 sequences remained, showing a removal of over 95% of genes from the gene abundance table. In contrast to Figure S3 where at the taxa level we did not see clusters between normotensive and hypertensive or masked hypertensive groups, at the biological pathway level these groups show distinct clustering (Figure 2D-E and Figure S9C-D). This supports that while the microbial taxa might be mostly similar between these groups, the gene pathways in each condition are different. These findings were also independent of clinic/location, sex, BMI and Australian dietary index scores (Figures S10 and S11). We identified 121 predicted microbiome gene pathways that were different as a result of the gut microbiota of hypertensive participants, 49 up-regulated and 72 down-regulated using PICRUSt2 (Table S6), and 69 pathways (31 up-regulated and 38 down-regulated) using Piphillin (Table S7). Particularly relevant were the down-regulation of genes involved in the sodium transport system and nitrite transporter, all relevant to hypertension and cardiovascular disease.

**Figure 2.**
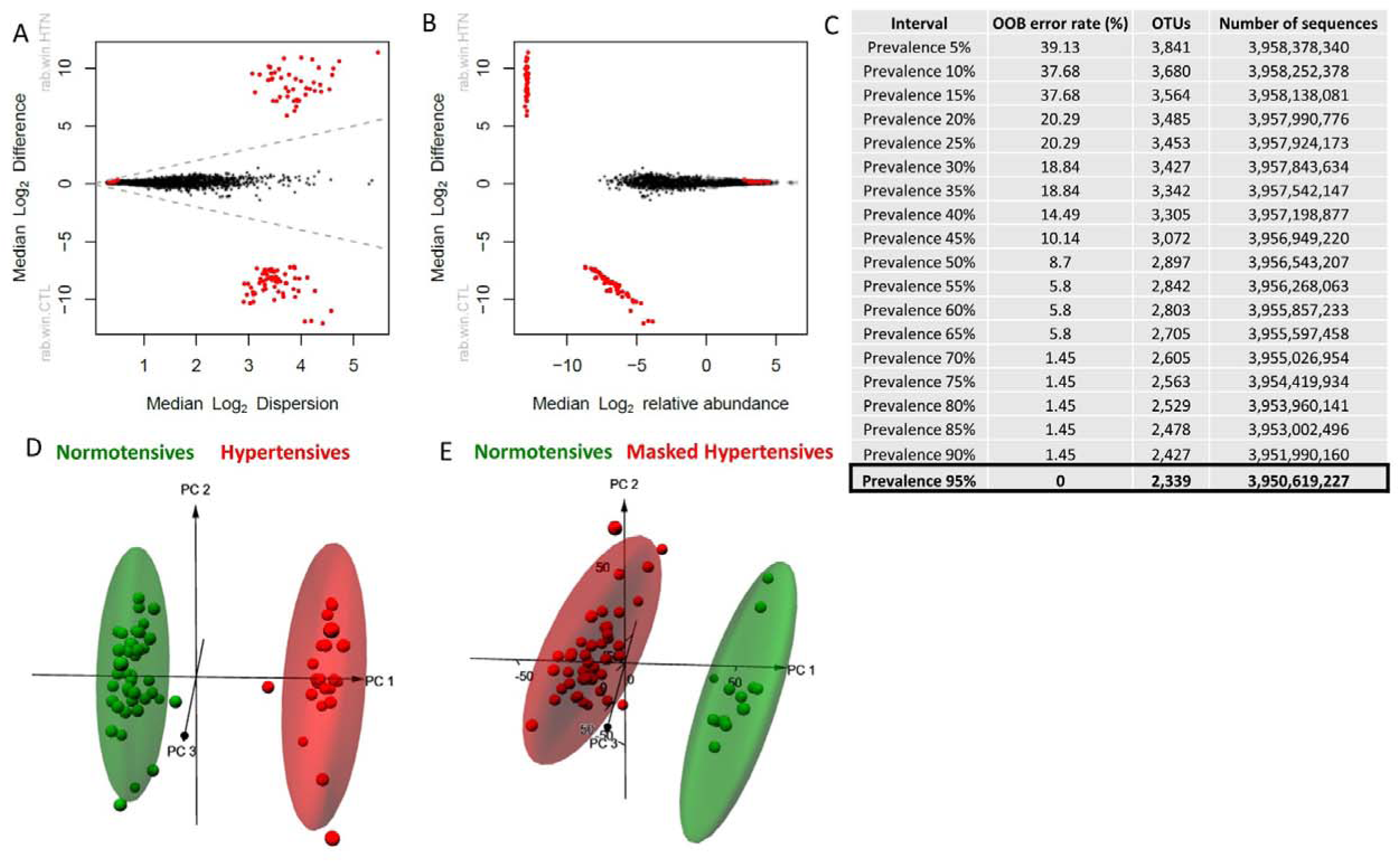
Machine-learning de-noised prevalence filtered gene data. **A.** Using the DADA2/PIME/ALDEx2 machine-learning pipeline described in Methods,^49^ the initial microbiome ASV dataset of 4,098 total raw data pathways was parsed into 5% intervals and ranked by out-of-bag (OOB) errors of signal-to-noise ratios giving minimized OBB error rate = 0 at the 95% prevalence level. This interval held 2,339 gene pathways. **B and C.** Mann-Whitney plot validating the 2,339 prevalent pathways (red points) of the 95% prevalence interval of (A). These pathways were subsequently used downstream for all the ensuing multivariate analyses comparing the groups. Points represent unique microbial gene functions that are differentially abundant at q<0.1 (red); abundant but non-differentially abundant (gray); rare and not differentially abundant (black). **D.** Principal component analysis (PCA) of prevalent microbial gene pathways using the data pool of (A) and (B), comparing essential hypertensive subjects (n=23) vs. normotensives (n=46). **E.** PCA comparing masked hypertensives (n=13) vs. normotensives (n=46). All PCA points are shown within 95% confidence interval ellipses. This figure shows data analysed with the PICRUSt2 server, while S8 shows data analysed with the Piphillin server.

Moreover, we observed a significant increase in acetate-CoA ligase (ADP-forming) in hypertensive participants (fold change 2.4, FDR-adjusted *P*= 1.97×10^-13^). This enzyme is responsible for the conversion of ATP, acetate and CoA into ADP, phosphate and acetyl-CoA, which is involved in the production of butyrate. This is relevant as acetate and butyrate are abundant SCFAs produced by the gut microbiota, and they are able to lower BP in experimental hypertension.^4, 20^ No other pathways and genes associated with SCFAs were differentially regulated in human hypertension in both Piphillin and PICRUSt2 servers (Figure S12).

### Short-chain fatty acids and receptors

To validate these findings, we then directly measured the levels of SCFAs. Hypertensive patients had higher levels of plasma, but not faecal (data not shown, all P>0.05), total levels of SCFAs compared to normotensive controls (*P*=0.04, Figure 3A). This was driven by higher levels of acetate (*P*=0.002, Figure 3B) and butyrate (*P*=0.0008, Figure 3C), but not propionate (*P* =0.32, Figure 3D). These associations were still significant after adjustment for age, BMI and sex (acetate: β= 0.012, *P*=0.05; butyrate: β= 0.519, *P*=0.006). Butyrate levels were positively correlated with 24-hour SBP (r=0.428, *P*=0.0009, Figure 3E) and DBP (r=0.453, *P*=0.0004, Figure 3F). We did not see any changes in SCFAs in patients with white coat hypertension, but those with masked hypertension also had higher levels of butyrate (mean±SEM: masked 6.42±0.6 vs others 4.88±0.3, *P*=0.027). Changes in plasma and faecal SCFAs were independent of dietary fibre, nut, legume, grain and vegetable intake (data not shown).

**Figure 3.**
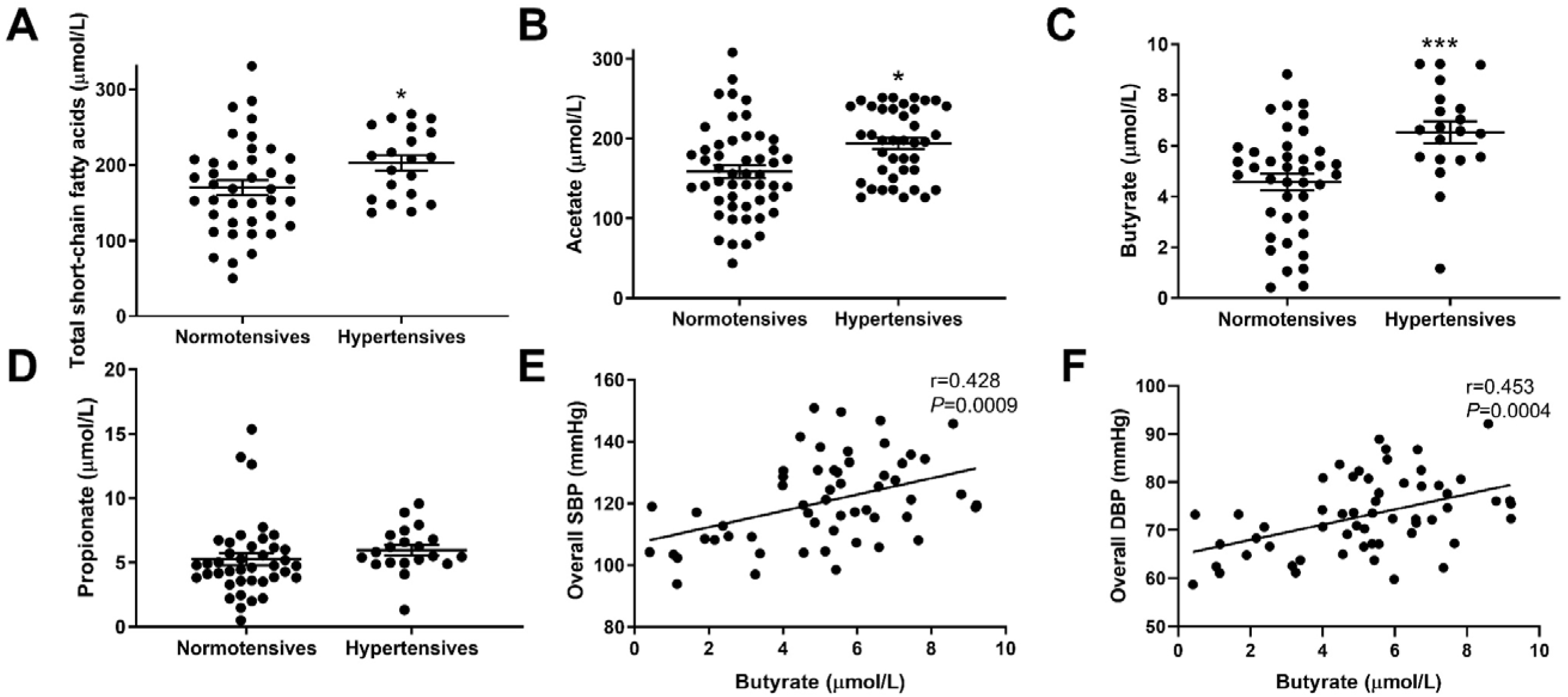
Plasma levels of gut microbial metabolites short-chain fatty acids (SCFAs) in association with hypertension. Showing A, total SCFAs, B, acetate, C, butyrate, and D, propionate. Showing mean ±standard error of mean. Correlations between butyrate and E, systolic blood pressure (SBP) and F, diastolic blood pressure (DBP). *indicates *P*<0.05, \*\*\**P*<0.001.

These findings, however, were in the opposite direction to what we expected since acetate and butyrate are able to lower BP in experimental hypertension.^4, 20^ We hypothesised that while hypertensive patients had higher levels of circulating SCFAs, they may lack the receptors that sense SCFAs. We then quantified the mRNA levels of SCFA-sensing receptors, GPR41, GPR43 and GPR109A, in white blood cells from these patients (Figure 4). We found that hypertensive patients had significantly lower levels of GPR43 (*FFAR2*) mRNA compared to normotensive controls (*P*<0.001). This was still significant after adjustment for age, BMI and sex (acetate: β= -12.3, P=0.004), where only GPR43 expression was still significantly associated with hypertension. GPR43 levels were also strongly negatively correlated with 24-hour SBP (ρ=-0.75, P<0.001) and DBP (ρ=-0.70, P<0.001). There were no changes in the expression of GPR41 and GPR019A. Our data suggest that untreated hypertensive patients have elevated circulating levels of acetate and butyrate, but lower levels of the SCFA-sensing receptor GPR43.

**Figure 4.**
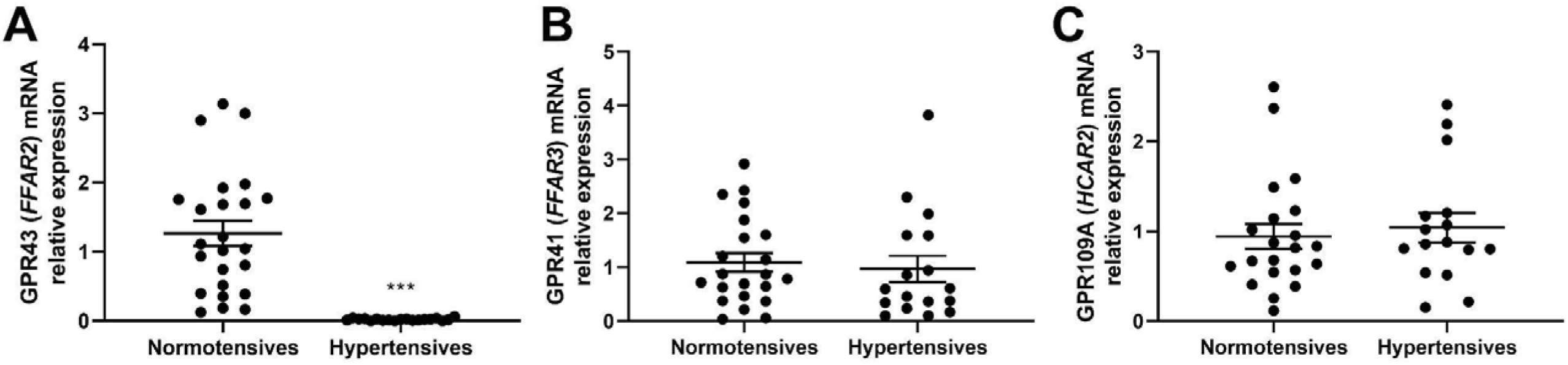
Relative expression of short-chain fatty acids (SCFAs)-sensing receptors, showing that A, *GPR43* mRNA is down-regulated in white blood cells of hypertensive participants, while there are no changes in B, *GPR41* and C, *GPR109A*. Showing mean ±standard error of mean. Sample size: n=25 normotensives and 22 hypertensives. ***indicates *P*<0.001.

## Discussion

Here we combined machine-learning, predicted gut microbiome sequencing, metabolite and receptor quantification with 24-hour BP monitoring from a multi-centre cohort. Our study uncovered that the gut microbiome of untreated essential, masked and white coat hypertensive subjects is similar to normotensive subjects in terms of diversity and composition, having only minor differences in taxa. It is, however, the predicted gut microbiota-derived gene pathways that are different across groups, particularly between normotensives and essential hypertensives. This was independent of traditional risk factors (e.g., age, sex and BMI) and microbiota risk factors (e.g., location, diet), and some of these changes were detected in the hosts’ circulation. One of the pathways identified involves the production of acetyl-CoA, which is relevant for the production of gut microbiota-derived SCFAs. Indeed, we found that hypertensive participants had higher plasma, but not faecal, levels of acetate and butyrate, but reduced levels of the main receptor that senses these metabolites, GPR43, in immune cells. Taken together, our study uncovered important metabolic gene pathways produced by the predicted human hypertensive gut microbiota and dysfunction of receptor targets that may drive an increase in BP and could form new therapeutic approaches for hypertension (summarised in Figure 5).

**Figure 5.**
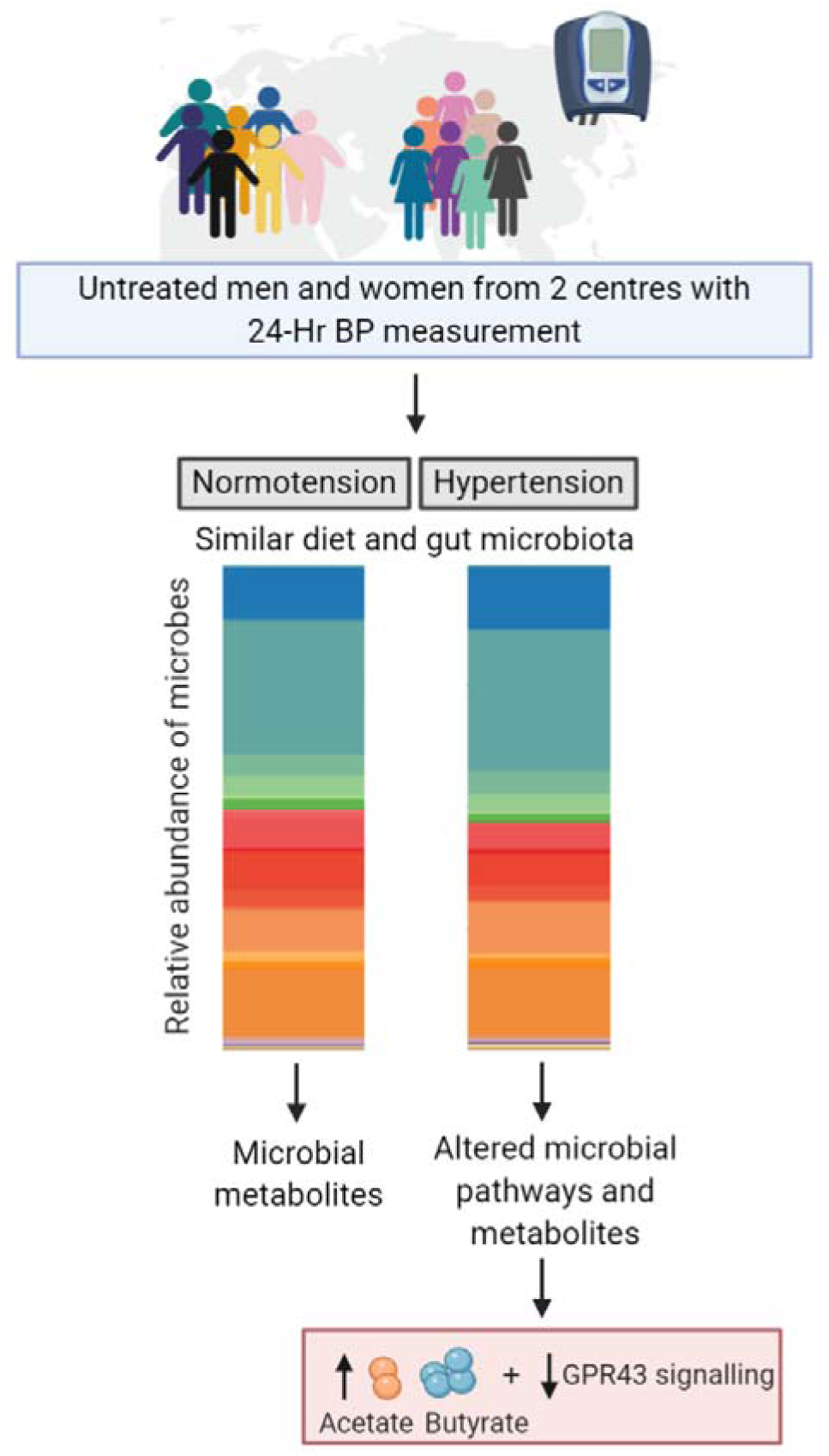
Summary of the study findings.

Contrary to some prior studies,^5, 7, 8, 10, 11^ but in accordance with others,^9, 12–14^ we did not observe a change in microbial α diversity between groups. Similarly, most studies have not been able to show a difference^10, 13^ or have not reported differences^5, 11, 14^ in microbial β diversity between people with normal and high BP. This may be explained by poor characterisation often with the use of office BP and inclusion of treated patients, amongst other factors.^14^ Indeed, to date only one study of 54 men recruited in one centre has analysed ABPM.^15^ Another explanation is the use of BP-lowering medication, which has been associated with changes in the gut microbiota,^15, 16^ as most studied besides one^5^ combined treated and untreated patients, while others classified treated patients with controlled BP as normotensive controls.^7, 9^ Albeit we did not identify β diversity changes between hypertensive groups, we still identified some differentially prevalent taxa. Particularly relevant are the lower levels of *Ruminococcus spp.* and *Eubacterium eligens* observed in both essential and masked hypertensive groups. *Ruminococcus spp.* are known as SCFA-producers,^55^ and have been previously associated with hypertension in other cohorts,^10, 11, 13, 14^ and recently in patients with heart failure with preserved ejection fraction (HFpEF),^56^ for which hypertension is a major risk factor for. Similarly, the less studied *Acidaminococcus spp* has also been reported.^5, 13^ Interestingly, hypertensive subjects had higher levels of taxa associated with degradation of mucin (mucosal-sugars), such as *Muribaculaceae spp.* and *Alistipes,*^57^ the latter being associated with epithelial inflammation.^58^

An alternative explanation to the lack of association with microbial diversity and different disease states has been recently introduced: diet may only modify the gut microbiota at the post-transcriptional (proteome) level, leading to altered protein and metabolite production while microbiota composition may remain the same.^59^ Our study provides the first human evidence that this may be the case for cardiovascular disease, particularly in the setting of untreated essential hypertension diagnosed with ABPM. We show that while α and β diversity remained unchanged, the predicted microbial gene networks associated with hypertension changed dramatically. Particularly relevant to hypertension, we identified that the human hypertension gut microbiota had higher expression of genes associated with sodium and nitrite transport, and acetate-CoA ligase, all relevant to the development of hypertension, but never before studied from a gut microbiota perspective. The role of intestinal bacteria, as opposed to oral bacteria,^60^ to the production of nitric oxide is not known, and was outside the scope of the present study. Similarly, it is still unknown whether bacterial sodium transport activity impacts on the host sodium absorption, but due to the role of the intestine in sodium absorption this could be a new direction to the field. We acknowledge as a limitation of our study that we did not quantify the levels of sodium in the faeces of our participants, because all the material had been used for SCFA and microbiome analyses. We also acknowledge that the difference in the microbial gene pathways detected are predictions and need to be validated with metatranscriptomics and proteomic analysis. This was outside the scope of the current study.

Gut microbial metabolites are key for microbial-host communication. One of the gene pathways we identified was an upregulation of acetate-CoA ligase in hypertensive subjects. Indeed, we found higher levels of acetate and butyrate, another SCFA that can also be a result of down-stream metabolism of acetate by acetate-CoA, in hypertensive subjects. Acetate and butyrate are sensed by three G-protein coupled receptors (GPCR), GPR43 being the major of these receptors. These SCFA-sensing receptors are highly expressed in immune cells,^52^ being one of the possible ways that microbiota-host communication takes place. Our study provides the first human evidence that GPR43 is differentially regulated in essential hypertension. One of the major findings of our study is that hypertensive subjects have a significantly lower expression of GPR43. Deficiency of GPR43 signalling results in immune dysfunction which drives a shift to a pro-inflammatory phenotype including recruitment of neutrophils and other immune cells.^61^ In a large follow-up study of 28,850 subjects, a greater peripheral neutrophil-to-lymphocyte ratio was found among hypertensive subjects proving inflammation exists in hypertensive subjects and at least involves an increase in neutrophils.^62^ Thus, the deficiency of GPR43 signalling may be (partially) responsible for the pro-inflammatory phenotype (increase in neutrophils) observed in hypertensive subjects. In the future, coupling the immune phenotype of subjects with metabolite-sensing GPCR expression is needed to further understand how SCFA-signalling affects the various immune cells. The increase we observed in SCFAs in hypertensive patients, however, remains unclear. The affinity of SCFA receptors has been shown to vary according to the concentrations of SCFAs,^63^ and the decrease in GPR43 could be a compensatory mechanism. This interaction between GPR43 and SCFAs in hypertension likely involves changes in the gastrointestinal tract, which is highly enriched with not only SCFA transporters but also with immune cells that highly express GPR43. Human intestinal tissue combined with colonic and plasma SCFA levels would be ideal to determine their relationship.

We acknowledge our study has some limitations, including the relatively small sample size. Albeit modest, our study took advantage of the only multi-site cohort published to date that has hypertension diagnosed by ABPM and includes both men and women, all untreated for BP-lowering medication. This is also the only cohort to date that contains detailed information regarding dietary intake as well as plasma and faecal metabolites, which allowed us to determine their relationship to the microbiota and BP, and show that our results are robust and valid across 2 independent recruitment sites. As a result of the detailed characterisation of this cohort including ABPM, our sample had similar power to identify microbial associations as some of the largest studies reported to date.^13^ However, due to the modest sample size, these results need to be independently validated. Ideally, a meta-analysis should be performed, however, this is impacted by the lack of studies with ABPM data.

## Perspectives

We identified that while predicted microbial diversity did not change in essential, masked and white coat hypertension diagnosed with 24-hour BP monitoring, there was a significant shift in microbial gene pathways involved in essential hypertension. Acetate-CoA ligase was up-regulated, coupled with higher levels of the SCFAs acetate and butyrate. Hypertensive subjects, however, had lower levels of the SCFA-sensing receptor GPR43 in immune cells, which may blunt their response to BP-lowering metabolites. This suggests that targeting the new pathways and the receptor identified here may represent new avenues in the search for mechanisms that control BP in hypertensive patients.

## Data Availability

The microbiome data underlying this article will be made available at the GenBank Nucleotide Database. The other data underlying this article will be shared on reasonable request to the corresponding author.

## Acknowledgements

We are grateful to Donna Vizi, Vivian Mak and Kaye Carter who assisted with blood collection. We also thank Professor Graham Giles of the Cancer Epidemiology & Intelligence Division, Cancer Council Victoria, for permission to use the Dietary Questionnaire for Epidemiological Studies (Version 3.2), Melbourne: Cancer Council Victoria, 1996.

## Author contribution

conceived and designed the study: FZM. Collected the data: FZM, SY, MC. Performed experiments: FM, MN, PG. Contributed data or analysis tools: FZM, MN, RR, BRS, GAH, PG, SY, MC, JM. Performed analyses: FZM, MN, PG, RR, BRS. Wrote the paper: FZM, MN, RR, RRM. Reviewed the paper: all authors. Secured funding: FZM, DMK.

## Data availability

The microbiome data underlying this article will be made available at the GenBank Nucleotide Database (under submission at the moment). The other data underlying this article will be shared on reasonable request to the corresponding author.

## Novelty and Significance

### 1. What is new?

- We performed the first predicted gut microbiome multi-site study of normotensive and untreated hypertensive participants, men and women, whose blood pressure was measured using ambulatory blood pressure monitoring.
- Normotensive and essential hypertensive groups could be differentiated based on predicted gut microbiome gene pathways and metabolites, but few differences were detected in terms of specific microbial types.
- Hypertensive patients exhibited higher levels of gut microbial-derived metabolites, but their immune cells expressed reduced levels of the main receptor that senses these metabolites.

### 2. What is relevant?

- This study addresses many gaps in the gut microbiota-hypertension literature including comparison across metropolitan and regional areas, men and women, all with 24-hour blood pressure measurements.
- The predicted gut microbiome was mostly similar between normotensive and essential hypertensive groups, but the predicted gut microbial gene pathways were different, suggesting major differences in the function of the microbiota.
- We identified that hypertensive subjects have a deficiency in a new target gene that senses gut microbiota-derived metabolites that lower blood pressure.

### 3. Summary

While predicted gut microbial diversity did not change in essential hypertension, there was a significant shift in microbial gene pathways, and an increase in the circulating levels of the SCFAs acetate and butyrate. Hypertensive subjects, however, had lower levels of the SCFA-sensing receptor GPR43, putatively blunting their response to BP-lowering metabolites.

## References

1. Tierney BT, Tan Y, Kostic AD, Patel CJ. Gene-level metagenomic architectures across diseases yield high-resolution microbiome diagnostic indicators. Nat Commun 2021(12):2907.

2. Muralitharan RR, Jama HA, Xie L, Peh A, Snelson M, Marques FZ. Microbial Peer Pressure: The Role of the Gut Microbiota in Hypertension and Its Complications. Hypertension 2020;76(6):1674–1687.

3. Marques FZ, Mackay CR, Kaye DM. Beyond gut feelings: how the gut microbiota regulates blood pressure. Nat Rev Cardiol 2018;15:20–32.

4. Kaye DM, Shihata W, Jama HA, Tsyganov K, Ziemann M, Kiriazis H, Horlock D, Vijay A, Giam B, Vinh A, Johnson C, Fiedler A, Donner D, Snelson M, Coughlan MT, Phillips S, Du XJ, El-Osta A, Drummond G, Lambert GW, Spector T, Valdes AM, Mackay CR, Marques FZ. Deficiency of Prebiotic Fibre and Insufficient Signalling Through Gut Metabolite Sensing Receptors Leads to Cardiovascular Disease. Circulation 2020(141):1393–1403.

5. Li J, Zhao F, Wang Y, Chen J, Tao J, Tian G, Wu S, Liu W, Cui Q, Geng B, Zhang W, Weldon R, Auguste K, Yang L, Liu X, Chen L, Yang X, Zhu B, Cai J. Gut microbiota dysbiosis contributes to the development of hypertension. Microbiome 2017;5(1):14.

6. Ferguson JF, Aden LA, Barbaro NR, Van Beusecum JP, Xiao L, Simmons AJ, Warden C, Pasic L, Himmel LE, Washington MK, Revetta FL, Zhao S, Kumaresan S, Scholz MB, Tang Z, Chen G, Reilly MP, Kirabo A. High dietary salt-induced dendritic cell activation underlies microbial dysbiosis-associated hypertension. JCI Insight 2019;5.

7. Yang T, Santisteban MM, Rodriguez V, Li E, Ahmari N, Carvajal JM, Zadeh M, Gong M, Qi Y, Zubcevic J, Sahay B, Pepine CJ, Raizada MK, Mohamadzadeh M. Gut dysbiosis is linked to hypertension. Hypertension 2015;65(6):1331–40.

8. Yan Q, Gu Y, Li X, Yang W, Jia L, Chen C, Han X, Huang Y, Zhao L, Li P, Fang Z, Zhou J, Guan X, Ding Y, Wang S, Khan M, Xin Y, Li S, Ma Y. Alterations of the Gut Microbiome in Hypertension. Front Cell Infect Microbiol 2017;7:381.

9. Kim S, Goel R, Kumar A, Qi Y, Lobaton G, Hosaka K, Mohammed M, Handberg EM, Richards EM, Pepine CJ, Raizada MK. Imbalance of gut microbiome and intestinal epithelial barrier dysfunction in patients with high blood pressure. Clin Sci (Lond) 2018;132(6):701–718.

10. Sun S, Lulla A, Sioda M, Winglee K, Wu MC, Jacobs DR, Jr., Shikany JM, Lloyd-Jones DM, Launer LJ, Fodor AA, Meyer KA. Gut Microbiota Composition and Blood Pressure. Hypertension 2019;73(5):998–1006.

11. Verhaar BJH, Collard D, Prodan A, Levels JHM, Zwinderman AH, Backhed F, Vogt L, Peters MJL, Muller M, Nieuwdorp M, van den Born BJH. Associations between gut microbiota, faecal short-chain fatty acids, and blood pressure across ethnic groups: the HELIUS study. Eur Heart J 2020.

12. Walejko JM, Kim S, Goel R, Handberg EM, Richards EM, Pepine CJ, Raizada MK. Gut microbiota and serum metabolite differences in African Americans and White Americans with high blood pressure. Int J Cardiol 2018;271:336–339.

13. Palmu J, Salosensaari A, Havulinna AS, Cheng S, Inouye M, Jain M, Salido RA, Sanders K, Brennan C, Humphrey GC, Sanders JG, Vartiainen E, Laatikainen T, Jousilahti P, Salomaa V, Knight R, Lahti L, Niiranen TJ. Association Between the Gut Microbiota and Blood Pressure in a Population Cohort of 6953 Individuals. J Am Heart Assoc 2020;9(15):e016641.

14. Huart J, Leenders J, Taminiau B, Descy J, Saint-Remy A, Daube G, Krzesinski JM, Melin P, de Tullio P, Jouret F. Gut Microbiota and Fecal Levels of Short-Chain Fatty Acids Differ Upon 24-Hour Blood Pressure Levels in Men. Hypertension 2019:HYPERTENSIONAHA11812588.

15. Zhernakova A, Kurilshikov A, Bonder MJ, Tigchelaar EF, Schirmer M, Vatanen T, Mujagic Z, Vila AV, Falony G, Vieira-Silva S, Wang J, Imhann F, Brandsma E, Jankipersadsing SA, Joossens M, Cenit MC, Deelen P, Swertz MA, LifeLines cohort s, Weersma RK, Feskens EJ, Netea MG, Gevers D, Jonkers D, Franke L, Aulchenko YS, Huttenhower C, Raes J, Hofker MH, Xavier RJ, Wijmenga C, Fu J. Population-based metagenomics analysis reveals markers for gut microbiome composition and diversity. Science 2016;352(6285):565–9.

16. Maier L, Pruteanu M, Kuhn M, Zeller G, Telzerow A, Anderson EE, Brochado AR, Fernandez KC, Dose H, Mori H, Patil KR, Bork P, Typas A. Extensive impact of non-antibiotic drugs on human gut bacteria. Nature 2018;555(7698):623–628.

17. David LA, Maurice CF, Carmody RN, Gootenberg DB, Button JE, Wolfe BE, Ling AV, Devlin AS, Varma Y, Fischbach MA, Biddinger SB, Dutton RJ, Turnbaugh PJ. Diet rapidly and reproducibly alters the human gut microbiome. Nature 2014;505(7484):559–63.

18. Park Y, Subar AF, Hollenbeck A, Schatzkin A. Dietary fiber intake and mortality in the NIH-AARP diet and health study. Arch Intern Med 2011;171(12):1061–8.

19. Reynolds A, Mann J, Cummings J, Winter N, Mete E, Te Morenga L. Carbohydrate quality and human health: a series of systematic reviews and meta-analyses. Lancet 2019;393(10170):434–445.

20. Marques FZ, Nelson E, Chu PY, Horlock D, Fiedler A, Ziemann M, Tan JK, Kuruppu S, Rajapakse NW, El-Osta A, Mackay CR, Kaye DM. High-Fiber Diet and Acetate Supplementation Change the Gut Microbiota and Prevent the Development of Hypertension and Heart Failure in Hypertensive Mice. Circulation 2017;135(10):964–977.

21. Bartolomaeus H, Balogh A, Yakoub M, Homann S, Marko L, Hoges S, Tsvetkov D, Krannich A, Wundersitz S, Avery EG, Haase N, Kraker K, Hering L, Maase M, Kusche-Vihrog K, Grandoch M, Fielitz J, Kempa S, Gollasch M, Zhumadilov Z, Kozhakhmetov S, Kushugulova A, Eckardt KU, Dechend R, Rump LC, Forslund SK, Muller DN, Stegbauer J, Wilck N. The Short-Chain Fatty Acid Propionate Protects from Hypertensive Cardiovascular Damage. Circulation 2019;139(11):1407–1421.

22. Parati G, Stergiou G, O’Brien E, Asmar R, Beilin L, Bilo G, Clement D, de la Sierra A, de Leeuw P, Dolan E, Fagard R, Graves J, Head GA, Imai Y, Kario K, Lurbe E, Mallion JM, Mancia G, Mengden T, Myers M, Ogedegbe G, Ohkubo T, Omboni S, Palatini P, Redon J, Ruilope LM, Shennan A, Staessen JA, vanMontfrans G, Verdecchia P, Waeber B, Wang J, Zanchetti A, Zhang Y, European Society of Hypertension Working Group on Blood Pressure M, Cardiovascular V. European Society of Hypertension practice guidelines for ambulatory blood pressure monitoring. J Hypertens 2014;32(7):1359–66.

23. Qiu C, Coughlin KB, Frederick IO, Sorensen TK, Williams MA. Dietary fiber intake in early pregnancy and risk of subsequent preeclampsia. Am J Hypertens 2008;21(8):903–9.

24. Zealand FSAN. AUSNUT 2007 - Australian Food Composition Tables In. Canberra, Australia; 2008.

25. Zealand FSAN. NUTTAB 2010 - Australian Food Composition Tables. In. Canberra, Australia; 2011.

26. Pretorius RA, Bodinier M, Prescott SL, Palmer DJ. Maternal Fiber Dietary Intakes during Pregnancy and Infant Allergic Disease. Nutrients 2019;11(8).

27. Zealand FSAN. AUSNUT 2011-13 Food Nutrient Database. In. Canberra, Australia; 2014.

28. Zealand FSAN. Data provided by food companies and organisations. In. Canberra, Australia; 2020.

29. Cashel K, English R, Lewis J. Composition of Food. Nutrition Section. . In. Canberra, Australia: Australian Government Publishing Services; 1989.

30. National Health and Medical Research Council. Australian Dietary Guidelines. In: NHMRC, (ed). Canberra, Australia; 2013.

31. Ward SJ, Coates AM, Hill AM. Application of an Australian Dietary Guideline Index to Weighed Food Records. Nutrients 2019;11(6).

32. McNaughton SA, Ball K, Crawford D, Mishra GD. An index of diet and eating patterns is a valid measure of diet quality in an Australian population. J Nutr 2008;138(1):86–93.

33. Ribeiro RV, Hirani V, Senior AM, Gosby AK, Cumming RG, Blyth FM, Naganathan V, Waite LM, Handelsman DJ, Kendig H, Seibel MJ, Simpson SJ, Stanaway F, Allman-Farinelli M, Le Couteur DG. Diet quality and its implications on the cardio-metabolic, physical and general health of older men: the Concord Health and Ageing in Men Project (CHAMP). Br J Nutr 2017;118(2):130–143.

34. Marques FZ, Jama HA, Tsyganov K, Gill PA, Rhys-Jones D, Muralitharan RR, Muir JG, Holmes AJ, Mackay CR. Guidelines for Transparency on Gut Microbiome Studies in Essential and Experimental Hypertension. Hypertension 2019;74:1279–1293.

35. Mirzayi C, Renson A, Zohra F, Elsafoury S, Geistlinger L, Kasselman L, Eckenrode K, van de Wijgert J, Loughman A, Marques FZ, Consortium; S, Consortium; GS, Society; MAaQC, Segata N, Huttenhower C, Dowd JB, Jones HE, L. W. Strengthening The Organization and Reporting of Microbiome Studies (STORMS): A Reporting Checklist for Human Microbiome Research. BioRvix 2021(Pre-print available at https://doi.org/10.1101/2020.06.24.167353).

36. Song SJ, Amir A, Metcalf JL, Amato KR, Xu ZZ, Humphrey G, Knight R. Preservation Methods Differ in Fecal Microbiome Stability, Affecting Suitability for Field Studies. mSystems 2016;1(3).

37. Caporaso JG, Lauber CL, Walters WA, Berg-Lyons D, Huntley J, Fierer N, Owens SM, Betley J, Fraser L, Bauer M, Gormley N, Gilbert JA, Smith G, Knight R. Ultra-high-throughput microbial community analysis on the Illumina HiSeq and MiSeq platforms. ISME J 2012;6(8):1621–4.

38. Bolyen E, Rideout JR, Dillon MR, Bokulich NA, Abnet CC, Al-Ghalith GA, Alexander H, Alm EJ, Arumugam M, Asnicar F, Bai Y, Bisanz JE, Bittinger K, Brejnrod A, Brislawn CJ, Brown CT, Callahan BJ, Caraballo-Rodriguez AM, Chase J, Cope EK, Da Silva R, Diener C, Dorrestein PC, Douglas GM, Durall DM, Duvallet C, Edwardson CF, Ernst M, Estaki M, Fouquier J, Gauglitz JM, Gibbons SM, Gibson DL, Gonzalez A, Gorlick K, Guo J, Hillmann B, Holmes S, Holste H, Huttenhower C, Huttley GA, Janssen S, Jarmusch AK, Jiang L, Kaehler BD, Kang KB, Keefe CR, Keim P, Kelley ST, Knights D, Koester I, Kosciolek T, Kreps J, Langille MGI, Lee J, Ley R, Liu YX, Loftfield E, Lozupone C, Maher M, Marotz C, Martin BD, McDonald D, McIver LJ, Melnik AV, Metcalf JL, Morgan SC, Morton JT, Naimey AT, Navas-Molina JA, Nothias LF, Orchanian SB, Pearson T, Peoples SL, Petras D, Preuss ML, Pruesse E, Rasmussen LB, Rivers A, Robeson MS, 2nd, Rosenthal P, Segata N, Shaffer M, Shiffer A, Sinha R, Song SJ, Spear JR, Swafford AD, Thompson LR, Torres PJ, Trinh P, Tripathi A, Turnbaugh PJ, Ul-Hasan S, van der Hooft JJJ, Vargas F, Vazquez-Baeza Y, Vogtmann E, von Hippel M, Walters W, Wan Y, Wang M, Warren J, Weber KC, Williamson CHD, Willis AD, Xu ZZ, Zaneveld JR, Zhang Y, Zhu Q, Knight R, Caporaso JG. Reproducible, interactive, scalable and extensible microbiome data science using QIIME 2. Nat Biotechnol 2019;37(8):852–857.

39. Callahan BJ, McMurdie PJ, Rosen MJ, Han AW, Johnson AJ, Holmes SP. DADA2: High-resolution sample inference from Illumina amplicon data. Nat Methods 2016;13(7):581–3.

40. Price MN, Dehal PS, Arkin AP. FastTree 2--approximately maximum-likelihood trees for large alignments. PLoS One 2010;5(3):e9490.

41. Katoh K, Misawa K, Kuma K, Miyata T. MAFFT: a novel method for rapid multiple sequence alignment based on fast Fourier transform. Nucleic Acids Res 2002;30(14):3059–66.

42. Bokulich NA, Kaehler BD, Rideout JR, Dillon M, Bolyen E, Knight R, Huttley GA, Gregory Caporaso J. Optimizing taxonomic classification of marker-gene amplicon sequences with QIIME 2’s q2-feature-classifier plugin. Microbiome 2018;6(1):90.

43. Bokulich NA, Dillon MR, Bolyen E, Kaehler BD, Huttley GA, Caporaso JG. q2-sample-classifier: machine-learning tools for microbiome classification and regression. J Open Res Softw 2018;3(30).

44. Segata N, Izard J, Waldron L, Gevers D, Miropolsky L, Garrett WS, Huttenhower C. Metagenomic biomarker discovery and explanation. Genome Biol 2011;12(6):R60.

45. Zakrzewski M, Proietti C, Ellis JJ, Hasan S, Brion MJ, Berger B, Krause L. Calypso: a user-friendly web-server for mining and visualizing microbiome-environment interactions. Bioinformatics 2017;33(5):782–783.

46. Iwai S, Weinmaier T, Schmidt BL, Albertson DG, Poloso NJ, Dabbagh K, DeSantis TZ. Piphillin: Improved Prediction of Metagenomic Content by Direct Inference from Human Microbiomes. PLoS One 2016;11(11):e0166104.

47. McMurdie PJ, Holmes S. phyloseq: an R package for reproducible interactive analysis and graphics of microbiome census data. PLoS One 2013;8(4):e61217.

48. Roesch LFW, Dobbler PT, Pylro VS, Kolaczkowski B, Drew JC, Triplett EW. pime: A package for discovery of novel differences among microbial communities. Mol Ecol Resour 2020;20(2):415–428.

49. Stevens BR, Roesch L, Thiago P, Russell JT, Pepine CJ, Holbert RC, Raizada MK, Triplett EW. Depression phenotype identified by using single nucleotide exact amplicon sequence variants of the human gut microbiome. Mol Psychiatry 2020.

50. Gill PA, van Zelm MC, Ffrench RA, Muir JG, Gibson PR. Successful elevation of circulating acetate and propionate by dietary modulation does not alter T-regulatory cell or cytokine profiles in healthy humans: a pilot study. Eur J Nutr 2019.

51. So D, Yao CK, Gill PA, Pillai N, Gibson PR, Muir JG. Screening dietary fibres for fermentation characteristics and metabolic profiles using a rapid in vitro approach: implications for irritable bowel syndrome. Br J Nutr 2020:1–11.

52. Muralitharan RR, Marques FZ. Diet-related gut microbial metabolites and sensing in hypertension. J Hum Hypertens 2021(35):162–169.

53. Motulsky HJ, Brown RE. Detecting outliers when fitting data with nonlinear regression - a new method based on robust nonlinear regression and the false discovery rate. BMC Bioinformatics 2006;7:123.

54. Wilck N, Matus MG, Kearney SM, Olesen SW, Forslund K, Bartolomaeus H, Haase S, Mahler A, Balogh A, Marko L, Vvedenskaya O, Kleiner FH, Tsvetkov D, Klug L, Costea PI, Sunagawa S, Maier L, Rakova N, Schatz V, Neubert P, Fratzer C, Krannich A, Gollasch M, Grohme DA, Corte-Real BF, Gerlach RG, Basic M, Typas A, Wu C, Titze JM, Jantsch J, Boschmann M, Dechend R, Kleinewietfeld M, Kempa S, Bork P, Linker RA, Alm EJ, Muller DN. Salt-responsive gut commensal modulates TH17 axis and disease. Nature 2017;551(7682):585–589.

55. Jama H, Beale A, Shihata WA, Marques FZ. The effect of diet on hypertensive pathology: is there a link via gut microbiota-driven immune-metabolism? Cardiovasc Res 2019;115(9):1435–1447.

56. Beale AL, O’Donnell JA, Nakai ME, Nanayakkara S, Vizi D, Carter K, Dean E, Ribeiro R, Yiallourou S, Carrington MJ, Marques FZ, Kaye DM. The gut microbiome of heart failure with preserved ejection fraction Journal of the American Heart Association 2021(In press).

57. Pereira FC, Wasmund K, Cobankovic I, Jehmlich N, Herbold CW, Lee KS, Sziranyi B, Vesely C, Decker T, Stocker R, Warth B, von Bergen M, Wagner M, Berry D. Rational design of a microbial consortium of mucosal sugar utilizers reduces Clostridiodes difficile colonization. Nat Commun 2020;11(1):5104.

58. Borton MA, Sabag-Daigle A, Wu J, Solden LM, O’Banion BS, Daly RA, Wolfe RA, Gonzalez JF, Wysocki VH, Ahmer BMM, Wrighton KC. Chemical and pathogen-induced inflammation disrupt the murine intestinal microbiome. Microbiome 2017;5(1):47.

59. Lobel L, Cao YG, Fenn K, Glickman JN, Garrett WS. Diet posttranslationally modifies the mouse gut microbial proteome to modulate renal function. Science 2020;369(6510):1518–1524.

60. Vanhatalo A, Blackwell JR, L’Heureux JE, Williams DW, Smith A, van der Giezen M, Winyard PG, Kelly J, Jones AM. Nitrate-responsive oral microbiome modulates nitric oxide homeostasis and blood pressure in humans. Free Radic Biol Med 2018;124:21–30.

61. Maslowski KM, Vieira AT, Ng A, Kranich J, Sierro F, Yu D, Schilter HC, Rolph MS, Mackay F, Artis D, Xavier RJ, Teixeira MM, Mackay CR. Regulation of inflammatory responses by gut microbiota and chemoattractant receptor GPR43. Nature 2009;461(7268):1282–6.

62. Liu X, Zhang Q, Wu H, Du H, Liu L, Shi H, Wang C, Xia Y, Guo X, Li C, Bao X, Su Q, Sun S, Wang X, Zhou M, Jia Q, Zhao H, Song K, Niu K. Blood Neutrophil to Lymphocyte Ratio as a Predictor of Hypertension. Am J Hypertens 2015;28(11):1339–46.

63. Le Poul E, Loison C, Struyf S, Springael JY, Lannoy V, Decobecq ME, Brezillon S, Dupriez V, Vassart G, Van Damme J, Parmentier M, Detheux M. Functional characterization of human receptors for short chain fatty acids and their role in polymorphonuclear cell activation. J Biol Chem 2003;278(28):25481–9.

