## Supplemental file for "Essential hypertension is associated with changes in gut microbial metabolic pathways: A multi-site analysis of ambulatory blood pressure"

**Tables**

**Table S1.** Strengthening The Organization and Reporting of Microbiome Studies (STORMS) reporting checklist.

| **Number** | **Item** | **Yes/No/NA** | **Comments or location in manuscript** |
| --- | --- | --- | --- |
| **Abstract** |  |  |  |
| 1 | Structured or Unstructured Abstract | Yes | Abstract |
| 1.1 | Study Design | Yes | Abstract |
| 1.2 | Sequencing methods | Yes | Abstract |
| 1.3 | Specimens | Yes | Abstract |
| **Introduction** |  |  |  |
| 2 | Background and Rationale | Yes | Page 5 |
| 2.1 | Hypotheses | Yes | Page 6 |
| **Methods** |  |  |  |
| 3 | Study Design | Yes | Page 6 (Participants and recruitment) |
| 3.1 | Participants | Yes | Page 6 (Participants and recruitment) |
| 3.2 | Geographic location | Yes | Page 6 (Participants and recruitment) |
| 3.3 | Relevant Dates | Yes | Page 6 (Participants and recruitment) |
| 3.4 | Eligibility criteria | Yes | Page 6 (Participants and recruitment) |
| 3.5 | Antibiotics Usage | Yes | Page 6 (Participants and recruitment) |
| 3.6 | Analytic sample size | Yes | Figure S2 |
| 3.7 | Longitudinal Studies | NA |  |
| 3.8 | Matching | NA |  |
| 3.9 | Ethics | Yes | Page 7 (Participants and recruitment) |
| 4 | Laboratory methods | Yes | Pages 8 and 9 (Faecal DNA extraction, library preparation and sequencing) |
| 4.1 | Specimen collection | Yes | Page 8 (Faecal DNA extraction, library preparation and sequencing) |
| 4.2 | Shipping | Yes | Page 8 (Faecal DNA extraction, library preparation and sequencing) |
| 4.3 | Storage | Yes | Page 8 (Faecal DNA extraction, library preparation and sequencing) |
| 4.4 | DNA extraction | Yes | Pages 8 and 9 (Faecal DNA extraction, library preparation and sequencing) |
| 4.5 | Human DNA sequence depletion or microbial DNA enrichment | NA |  |
| 4.6 | Primer selection | Yes | Pages 8 and 9 (Faecal DNA extraction, library preparation and sequencing) |
| 4.7 | Positive Controls | Yes | Page 9 (Faecal DNA extraction, library preparation and sequencing) |
| 4.8 | Negative Controls | Yes | Page 9 (Faecal DNA extraction, library preparation and sequencing) |
| 4.9 | Contaminant mitigation and identification | Yes | Page 9 (Faecal DNA extraction, library preparation and sequencing) |
| 4.1 | Replication | Yes | Page 9 (Faecal DNA extraction, library preparation and sequencing) |
| 4.11 | Sequencing strategy | Yes | Page 8 (Faecal DNA extraction, library preparation and sequencing) |
| 4.12 | Sequencing methods | Yes | Page 9 (Faecal DNA extraction, library preparation and sequencing) |
| 4.13 | Batch effects | NA | Not relevant as all samples were extracted and sequenced together |
| 4.14 | Metatranscriptomics | NA |  |
| 4.15 | Metaproteomics | NA |  |
| 4.16 | Metabolomics | Yes | Page 11 (Short-chain fatty acids measurement) |
| 5 | Data sources/  measurement | Yes | Pages 7 and 8 (Blood pressure measurement and hypertension diagnosis, and Food frequency questionnaire) |
| 6 | Research design for causal inference | Yes | Page 12 (Statistical analyses) |
| 6.1 | Selection bias | NA |  |
| 7 | Bioinformatic and Statistical Methods | Yes | Pages 9-11 (Bioinformatic analyses of gut microbiomes) |
| 7.1 | Quality Control | Yes | Pages 9-11 (Bioinformatic analyses of gut microbiomes) |
| 7.2 | Sequence analysis | Yes | Pages 9-11 (Bioinformatic analyses of gut microbiomes) |
| 7.3 | Statistical methods | Yes | Pages 9-12 (Bioinformatic analyses of gut microbiomes, and Statistical analyses) |
| 7.4 | Longitudinal analysis | NA |  |
| 7.5 | Subgroup analysis | Yes | Pages 9-11 (Bioinformatic analyses of gut microbiomes) |
| 7.6 | Missing data | NA |  |
| 7.7 | Sensitivity analyses | Yes | Page 12 (Statistical analyses) |
| 7.8 | Findings | Yes | Pages 9-12 (Bioinformatic analyses of gut microbiomes, and Statistical analyses) |
| 7.9 | Software | Yes | Pages 9-12 (Bioinformatic analyses of gut microbiomes, and Statistical analyses) |
| 8 | Reproducible research | Yes | Pages 9-11 (Bioinformatic analyses of gut microbiomes) |
| 8.1 | Raw data access | Yes | Pages 9-11 (Bioinformatic analyses of gut microbiomes) |
| 8.2 | Processed data access | No | We have provided access to all raw data. |
| 8.3 | Participant data access | No | Individual participant data cannot be provided. |
| 8.4 | Source code access | Yes | <https://gitlab.erc.monash.edu.au/marqueslab> |
| 8.5 | Full results | No | Values have been inputed into Tables in the supplementary files as they are easier to read. |
| **Results** |  |  |  |
| 9 | Descriptive data | Yes | Pages 12 and 3 (Participants’ characteristics, and Dietary food intake) |
| 10 | Microbiome data | Yes | Pages 13-15 (Gut microbiome) |
| 10.1 | Taxonomy | Yes | Pages 13-15 (Gut microbiome) |
| 10.2 | Differential abundance | Yes | Pages 13-15 (Gut microbiome) |
| 10.3 | Other data types | Yes | Pages 13-15 (Gut microbiome) |
| 10.4 | Other statistical analysis | Yes | Page 16 (Short-chain fatty acids and receptors) |
| **Discussion** |  |  |  |
| 11 | Key results | Yes | Page 17 (first paragraph) |
| 12 | Interpretation | Yes | Pages 17-20 |
| 13 | Limitations | Yes | Page 20 |
| 13.1 | Bias | No |  |
| 13.2 | Generalizability | Yes | Pages 20 and 21 (Perspectives) |
| 14 | Ongoing/future work | Yes | Pages 20 and 21 (Perspectives) |
| **Other information** |  |  |  |
| 15 | Funding | Yes | Page 21 (Sources of funding) |
| 15.1 | Acknowledgements | Yes | Page 21 (Acknowledgements) |
| 15.2 | Conflicts of Interest | Yes | Page 21 (Disclosures) |
| 16 | Supplements | Yes | Attached, journal will link them |
| 17 | Supplementary data | Yes | Attached, journal will link them |

**Table S2.** TaqMan assays used in the study.

| **Genes** | **Assay ID** |
| --- | --- |
| *FFAR3* (GPR41) | Hs02519193_g1 |
| *FFAR2* (GPR43) | Hs00271142_s1 |
| *HCAR2* (GPR109A) | Hs02341584_s1 |
| *ACTB* (housekeeping gene) | Hs01060665_g1 |
| *GAPDH* (housekeeping gene) | Hs02786624_g1 |

**Table S3.** Dietary variables analysed in relationship with hypertension.

| **Food type in grams (mean±SEM)** | **Normotensives (n=46)** | **Hypertensives (n=23)** | ***P*-value** |
| --- | --- | --- | --- |
| Total dietary fibre | 23.54±1.3 | 22.9±2.0 | 0.781 |
| Total resistant starches | 2.09±0.1 | 2.07±0.2 | 0.947 |
| Total insoluble fibre | 8.44±0.5 | 8.02±0.8 | 0.672 |
| Total soluble fibre | 6.08±0.3 | 5.78±0.5 | 0.606 |
| Nuts | 18.15±3.3 | 14.5±2.6 | 0.384 |
| Vegetables | 249.0±20.6 | 228.8±19.4 | 0.479 |
| Dark green vegetables | 34.05±5.4 | 26.91±5.1 | 0.341 |
| Starchy vegetables | 32.69±3.8 | 30.61±4.2 | 0.716 |
| Legumes | 49.11±8.2 | 60.9±14 | 0.471 |
| Dry fruit | 11.26±2.8 | 6.91±2.1 | 0.218 |
| Fresh fruit | 266.9±25.3 | 234.9±33.3 | 0.448 |
| Grain source of fibre | 121.6±14.7 | 114.1±11.2 | 0.349 |
| Grains total | 213.0±18.0 | 216.4±15.6 | 0.885 |
| Grains low in fibre | 91.42±8.0 | 112.3±11.3 | 0.139 |

**Table S4**. Alpha diversity comparison between normotensive and hypertensive groups.

| **α diversity metric (mean±SEM)** | **Normotensives (n=46)** | **Hypertensives (n=23)** | ***P*-value** |
| --- | --- | --- | --- |
| Observed OTUs | 224.1±9.6 | 234.8±14.4 | 0.541 |
| Chao1 | 227.4±9.7 | 236.3±14.5 | 0.614 |
| Shannon | 3.7±0.1 | 3.8±0.1 | 0.972 |
| Simpson | 0.932±0.008 | 0.934±0.013 | 0.907 |
| Inverse Simpson | 21.6±1.7 | 21.9±2.1 | 0.906 |

**Legend**: OTUs, operational taxonomic units.

**Table S5.** Taxa correlated with 24-hour ambulatory blood pressure with P<0.0.5.

| **Genus (or species, when classified)** | **r** | ***P*-value** | **FDR** | **Blood pressure type** |
| --- | --- | --- | --- | --- |
| *Butyricicoccus* | 0.36 | 0.002668 | 0.48 | 24h SBP |
| ***Acidaminococcus*** | 0.34 | 0.005085 | 0.75 | 24h DBP |
| ***Ruminococcus*** | -0.32 | 0.008144 | 0.54 | 24h SBP |
| *Eubacterium hallii* | -0.31 | 0.009311 | 0.54 | 24h SBP |
| *Anaerotruncus* | -0.3 | 0.01256 | 0.54 | 24h SBP |
| ***Eubacterium eligens*** | -0.3 | 0.012968 | 0.75 | 24h DBP |
| *Anaerotruncus* | -0.3 | 0.014105 | 0.75 | 24h DBP |
| ***Acidaminococcus*** | 0.28 | 0.018562 | 0.55 | 24h SBP |
| ***Eubacterium eligens*** | -0.28 | 0.021752 | 0.56 | 24h SBP |
| ***Ruminococcus*** | -0.26 | 0.030806 | 0.82 | 24h DBP |
| *GCA900066225* | -0.26 | 0.030849 | 0.82 | 24h DBP |
| *Butyricicoccus* | -0.25 | 0.039161 | 0.7 | 24h SBP |
| *Anaerofilum* | -0.25 | 0.03979 | 0.82 | 24h DBP |
| *Butyricicoccus* | 0.24 | 0.044823 | 0.82 | 24h DBP |
| *Caproiciproducens* | -0.24 | 0.045366 | 0.82 | 24h DBP |
| *Catenibacterium* | -0.24 | 0.045394 | 0.7 | 24h SBP |
| *Ruminococcus torques* | -0.24 | 0.046772 | 0.7 | 24h SBP |
| *GCA900066225* | -0.24 | 0.047087 | 0.7 | 24h SBP |

In bold taxa also identified in the LDA analysis shown in Figure 1.

**Table S6.** Pathways differentially regulated in essential hypertension according to the PICRUSt2 server (effect size < or >50% change and q<0.05).

| **Taxcodes** | **Gene pathways names** | **Effect** |
| --- | --- | --- |
| K04027 | ethanolamine_utilization_protein_EutM | -3.51 |
| K00261 | glutamate_dehydrogenase_NAD_P__ | 2.96 |
| K01906 | 6_carboxyhexanoate_CoA_ligase | 2.80 |
| K03490 | AraC_family_transcriptional_regulator_dual_regulator_of_chb_operon | -3.42 |
| K07504 | predicted_type_IV_restriction_endonuclease | -3.30 |
| K06413 | stage_V_sporulation_protein_K | 2.11 |
| K00856 | adenosine_kinase | -3.07 |
| K03191 | acid_activated_urea_channel | -3.12 |
| K01250 | pyrimidine_specific_ribonucleoside_hydrolase | -2.99 |
| K03457 | nucleobasecation_symporter_1_NCS1_family | -3.22 |
| K07486 | transposase | 3.07 |
| K02784 | phosphocarrier_protein_HPr | -2.61 |
| K02083 | allantoate_deiminase | -2.90 |
| K07275 | outer_membrane_protein | 2.85 |
| K05303 | O_methyltransferase | -3.02 |
| K00219 | 2_4_dienoyl_CoA_reductase_NADPH2_ | -2.91 |
| K07337 | penicillin_binding_protein_activator | 2.95 |
| K07013 | uncharacterized_protein | -2.80 |
| K05773 | tungstate_transport_system_permease_protein | -2.35 |
| K07442 | tRNA_adenine57_N1_adenine58_N1_methyltransferase_catalytic_subunit | -2.63 |
| K05772 | tungstate_transport_system_substrate_binding_protein | -2.33 |
| K06951 | uncharacterized_protein | -2.53 |
| K07096 | uncharacterized_protein | -2.50 |
| K08296 | phosphohistidine_phosphatase | 2.25 |
| K06857 | tungstate_transport_system_ATP_binding_protein | -2.36 |
| K00437 | hydB | -2.51 |
| K06195 | ApaG_protein | 2.48 |
| K00534 | ferredoxin_hydrogenase_small_subunit | -2.68 |
| K07276 | uncharacterized_protein | 2.01 |
| K07640 | two_component_system_OmpR_family_sensor_histidine_kinase_CpxA | -2.68 |
| K08084 | type_IV_fimbrial_biogenesis_protein_FimT | -2.70 |
| K07062 | toxin_FitB | -2.58 |
| K00381 | sulfite_reductase_NADPH_hemoprotein_beta_component | 1.90 |
| K02173 | putative_kinase | -2.43 |
| K03197 | type_IV_secretion_system_protein_VirB2 | -2.47 |
| K03399 | cobalt_precorrin_7_C5_methyltransferase | -2.28 |
| K05685 | macrolide_transport_system_ATP_binding_permease_protein | -2.39 |
| K02191 | cobalt_precorrin_6B_C15_methyltransferase | -2.25 |
| K07662 | two_component_system_OmpR_family_response_regulator_CpxR | -2.30 |
| K03297 | small_multidrug_resistance_pump | -2.67 |
| K03851 | taurine_pyruvate_aminotransferase | -2.34 |
| K08173 | MFS_transporter_MHS_family_metaboliteH_symporter | -2.27 |
| K07048 | phosphotriesterase_related_protein | -2.47 |
| K06605 | myo_inositol_catabolism_protein_IolH | 2.73 |
| K03093 | RNA_polymerase_sigma_factor | -2.08 |
| K07046 | L_fuconolactonase | -2.24 |
| K00632 | acetyl_CoA_acyltransferase | 2.20 |
| K00899 | 5_methylthioribose_kinase | 2.44 |
| K06285 | transcription_attenuation_protein_tryptophan_RNA_binding_attenuator_protein_ | 2.10 |
| K02783 | glucitol_sorbitol_PTS_system_EIIC_component | -2.59 |
| K05346 | deoxyribonucleoside_regulator | 2.58 |
| K09136 | ribosomal_protein_S12_methylthiotransferase_accessory_factor | 2.60 |
| K02781 | glucitol_sorbitol_PTS_system_EIIA_component | -2.81 |
| K00068 | sorbitol_6_phosphate_2_dehydrogenase | -2.56 |
| K02549 | o_succinylbenzoate_synthase | -1.87 |
| K00045 | mannitol_2_dehydrogenase | 2.87 |
| K00532 | ferredoxin_hydrogenase | 2.41 |
| K05299 | formate_dehydrogenase_NADP_alpha_subunit | -2.29 |
| K07710 | two_component_system_NtrC_family_sensor_histidine_kinase_AtoS | -2.25 |
| K01640 | hydroxymethylglutaryl_CoA_lyase | 1.82 |
| K01669 | deoxyribodipyrimidine_photo_lyase | 1.80 |
| K07821 | trimethylamine_N_oxide_reductase_cytochrome_c_cytochrome_c_type_subunit_TorY | -2.40 |
| K08680 | 2_succinyl_6_hydroxy_2_4_cyclohexadiene_1_carboxylate_synthase | -1.83 |
| K06426 | small_acid_soluble_spore_protein_I_minor_ | 2.58 |
| K01029 | 3_oxoacid_CoA_transferase_subunit_B | 2.00 |
| K09459 | phosphonopyruvate_decarboxylase | -2.80 |
| K00823 | 4_aminobutyrate_aminotransferase | -2.04 |
| K02552 | menaquinone_specific_isochorismate_synthase | -1.89 |
| K03651 | 3_5_cyclic_AMP_phosphodiesterase | -2.27 |
| K01841 | phosphoenolpyruvate_phosphomutase | -2.67 |
| K02168 | choline_glycine_proline_betaine_transport_protein | -2.25 |
| K05774 | ribose_1_5_bisphosphokinase | -2.06 |
| K01066 | acetyl_esterase | -2.10 |
| K06175 | tRNA_pseudouridine65_synthase | 2.20 |
| K02815 | sorbose_PTS_system_EIID_component | -2.21 |
| K02847 | O_antigen_ligase | 2.15 |
| K06155 | Gnt_I_system_high_affinity_gluconate_transporter | 2.65 |
| K03532 | trimethylamine_N_oxide_reductase_cytochrome_c_cytochrome_c_type_subunit_TorC | -2.25 |
| K03762 | MFS_transporter_MHS_family_proline_betaine_transporter | 1.95 |
| K00383 | glutathione_reductase_NADPH_ | 1.80 |
| K07063 | uncharacterized_protein | 2.42 |
| K09472 | 4_gamma_glutamylamino_butanal_dehydrogenase | -2.05 |
| K04748 | nitric_oxide_reductase_NorQ_protein | 2.62 |
| K02196 | heme_exporter_protein_D | 2.03 |
| K07344 | type_IV_secretion_system_protein_TrbL | -2.14 |
| K03198 | type_IV_secretion_system_protein_VirB3 | -2.22 |
| K07305 | peptide_methionine_R_S_oxide_reductase | 2.02 |
| K02814 | sorbose_PTS_system_EIIC_component | -2.22 |
| K03221 | type_III_secretion_protein_F | -2.10 |
| K00228 | coproporphyrinogen_III_oxidase | 2.10 |
| K08094 | 6_phospho_3_hexuloisomerase | 1.83 |
| K06165 | alpha_D_ribose_1_methylphosphonate_5_triphosphate_synthase_subunit_PhnH | -2.08 |
| K03224 | ATP_synthase_in_type_III_secretion_protein_N | -2.10 |
| K07168 | CBS_domain_containing_membrane_protein | -2.21 |
| K02077 | zinc_manganese_transport_system_substrate_binding_protein | 1.96 |
| K05835 | threonine_efflux_protein | 1.80 |
| K02594 | homocitrate_synthase_NifV | -2.00 |
| K06917 | tRNA_2_selenouridine_synthase | -1.89 |
| K03342 | para_aminobenzoate_synthetase_4_amino_4_deoxychorismate_lyase | 2.01 |
| K03228 | type_III_secretion_protein_T | -2.05 |
| K03219 | type_III_secretion_protein_C | -2.01 |
| K05844 | ribosomal_protein_S6_L_glutamate_ligase | 1.84 |
| K03225 | type_III_secretion_protein_Q | -2.02 |
| K03226 | type_III_secretion_protein_R | -2.16 |
| K04058 | type_III_secretion_protein_W | -2.07 |
| K03222 | type_III_secretion_protein_J | -2.06 |
| K08484 | phosphotransferase_system_enzyme_I_PtsP | 1.78 |
| K07243 | high_affinity_iron_transporter | 1.94 |
| K01753 | D_serine_dehydratase | 1.99 |
| K04025 | ethanolamine_utilization_protein_EutK | -2.03 |
| K02786 | lactose_PTS_system_EIIA_component | 1.73 |
| K03229 | type_III_secretion_protein_U | -2.12 |
| K02291 | 15_cis_phytoene_synthase | 1.65 |
| K03230 | type_III_secretion_protein_V | -2.10 |
| K03697 | ATP_dependent_Clp_protease_ATP_binding_subunit_ClpE | 1.52 |
| K00380 | sulfite_reductase_NADPH_flavoprotein_alpha_component | 1.77 |
| K06191 | glutaredoxin_like_protein_NrdH | 1.69 |
| K06975 | uncharacterized_protein | 0.60 |
| K01620 | threonine_aldolase | 0.51 |
| K03575 | A_G_specific_adenine_glycosylase | 0.49 |
| K03284 | magnesium_transporter | 0.42 |

**Table S7.** Pathways differentially regulated in essential hypertension according to the Piphillin server (effect size < or >50% change and q<0.05).

| **Taxcodes** | **Gene pathways names** | **Effect** |
| --- | --- | --- |
| K17234 | arabinosaccharide transport system substrate binding protein 1 | -2.83 |
| K17235 | arabinosaccharide transport system permease protein 1 | -2.65 |
| K01308 | g D glutamyl meso diaminopimelate peptidase 1 | -2.60 |
| K17236 | arabinosaccharide transport system permease protein 2 | -2.57 |
| K10200 | N acetylglucosamine transport system substrate binding protein 1 | -2.45 |
| K00936 | two-component system sensor, histidine kinase PdtaS 1 | -2.43 |
| K10201 | N acetylglucosamine transport system permease protein 1 | -2.40 |
| K10202 | N acetylglucosamine transport system permease protein 2 | -2.33 |
| K07459 | putative ATP-dependent endonuclease of the OLD family 1 | -2.28 |
| K07321 | Codehydrogenase maturation factor 1 | -2.26 |
| K00042 | 2-hydroxy-3-oxopropionate reductase 1 | -2.19 |
| K04784 | yersiniabactin non-ribosomal peptide synthetase 1 | -2.15 |
| K05795 | tellurium resistance protein TerD 1 | -2.15 |
| K04783 | yersiniabactin salicyl AM Pligase 1 | -2.10 |
| K05341 | amylosucrase 1 | -2.05 |
| K06331 | spore coat proteinI 1 | -2.03 |
| K00786 | beta 1 6 galactosyltransferase 1 | -2.01 |
| K19504 | glucose lysine 6 phosphate deglycase 1 | -1.98 |
| K09696 | sodium transport system permease protein 1 | -1.94 |
| K09697 | sodium transport system ATP binding protein 1 | -1.93 |
| K19165 | antitoxin Phd 1 | -1.88 |
| K09703 | uncharacterized protein 223 | -1.87 |
| K01060 | cephalosporin C-deacetylase 1 | -1.77 |
| K18344 | two-component system, Omp R-family response regulator VanR 1 | -1.77 |
| K16139 | glucuronide carrier-protein 1 | -1.74 |
| K02598 | nitrite transporter 1 | -1.70 |
| K03486 | GntR-family transcriptional regulator trehalose operon transcriptional repressor 1 | -1.68 |
| K14205 | phosphatidyl glycerollysyl transferase 1 | -1.67 |
| K02977 | ubiquitin small subunit ribosomal protein S27Ae 1 | -1.66 |
| K00632 | acetyl CoA-acyltransferase 1 | -1.63 |
| K07130 | arylformamidase 2 | -1.60 |
| K00004 | (R,R)-butanediol dehydrogenase meso-butanediol dehydrogenase diacetylreductase 1 | -1.58 |
| K09685 | purine operon repressor 1 | -1.57 |
| K16907 | fluoroquinolone transport system ATP binding protein 1 | -1.53 |
| K22103 | DeoR family transcriptional regulator carbon catabolite repression regulator 1 | -1.52 |
| K07406 | alpha galactosidase 2 | -1.52 |
| K08981 | putative membrane protein 12 | -1.50 |
| K05363 | serine/alanine adding enzyme 1 | -1.50 |
| K14347 | solute carrier family 10 sodium bile acid cotransporte R member7 1 | 1.50 |
| K10806 | acyl CoA-thioesterase YciA 1 | 1.51 |
| K06181 | 23S rRNA pseudouridine^2457^ synthase 1 | 1.51 |
| K03652 | DNA 3-methyl-adenine glycosylase 1 | 1.52 |
| K07102 | N acetylmuramate1 kinase 1 | 1.53 |
| K14682 | amino acid N acetyltransferase 3 | 1.53 |
| K08300 | ribonucleaseE 1 | 1.54 |
| K06191 | glutaredoxin likeprotein NrdH 1 | 1.54 |
| K03117 | sec-independent protein translocase protein TatB 1 | 1.55 |
| K01792 | glucose 6 phosphate1 epimerase 1 | 1.57 |
| K01598 | phosphopantothenoylcysteine decarboxylase 1 | 1.62 |
| K00324 | H translocating NAD P-transhydrogenase subunit alpha 1 | 1.62 |
| K21977 | phosphopantothenate cysteine-ligase CTP 1 | 1.63 |
| K21030 | D-ribitol-5-phosphate cytidylyltransferase 1 | 1.64 |
| K01066 | acetylesterase 1 | 1.65 |
| K01708 | galactarate dehydratase 1 | 1.65 |
| K07219 | putative molybdopterin biosynthesis protein 1 | 1.66 |
| K03579 | ATP dependent-helicase HrpB 1 | 1.72 |
| K07318 | adenine specificDNA methyltransferase 3 | 1.73 |
| K01239 | purine nucleosidase 1 | 1.76 |
| K05939 | acyl-acyl carrier protein phospholipid O-acyltransferase long-chain fatty acid acyl carrier protein ligase 1 | 1.76 |
| K00184 | dimethyl sulfoxide reductase iron sulfurs ubunit 1 | 1.78 |
| K02584 | Nif specific-regulatory protein 1 | 1.79 |
| K12506 | 2-C-methyl-D-erythritol 4-phosphatecytidylyl transferase 2-C-methyl D erythritol 2-4-cyclodiphosphate synthase 1 | 1.81 |
| K00185 | dimethyl sulfoxide reductase membrane subunit 1 | 1.83 |
| K09007 | GTP cyclohydrolaseI B 1 | 1.86 |
| K03684 | ribonucleaseD 1 | 1.94 |
| K18816 | aminoglycoside 6-N-acetyltransferaseI 2 | 1.97 |
| K11934 | outer membrane protein-X 1 | 1.99 |
| K07304 | peptide methionine (S)-S oxide reductase 1 | 2.35 |
| K01905 | acetate CoA-ligase ADP forming subunit alpha 1 | 2.38 |

**Figures**

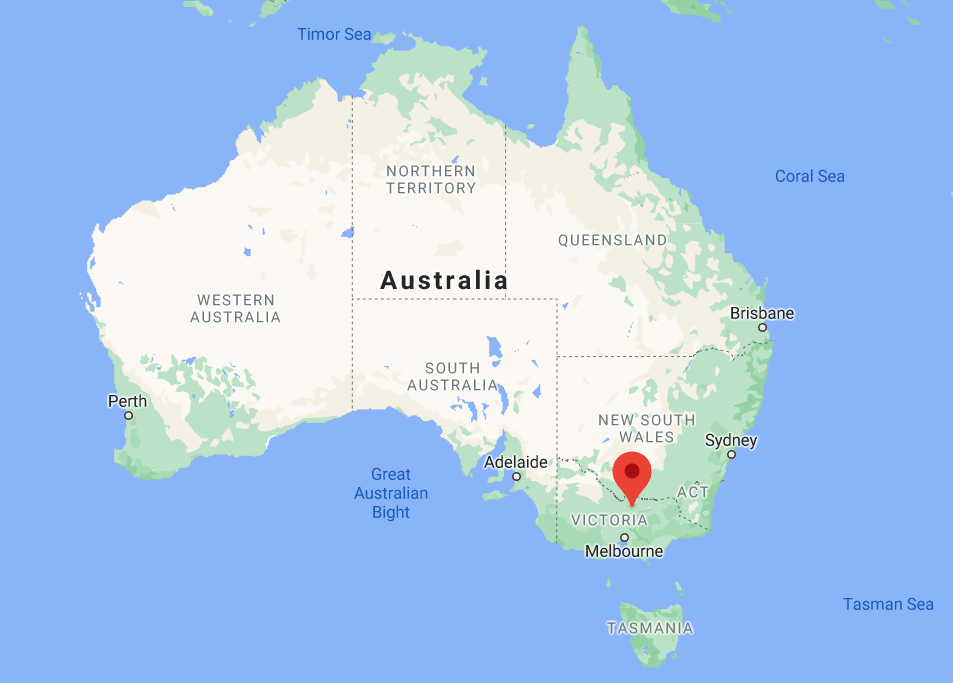

**Figure S1.** Map showing the two sites used for recruitment, Melbourne and Shepparton (marked with the red pin).

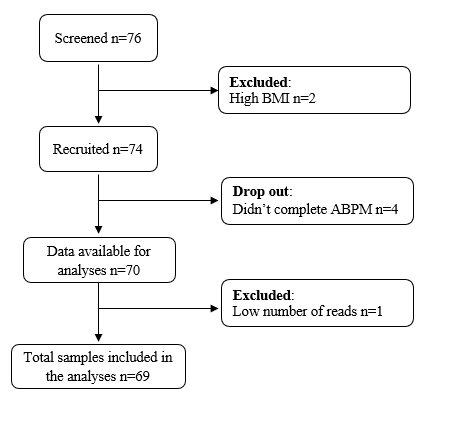

**Figure S2**. Strengthening The Organization and Reporting of Microbiome Studies (STORMS) flowchart. Legend: ABPM, ambulatory blood pressure monitoring; BMI, body mass index.

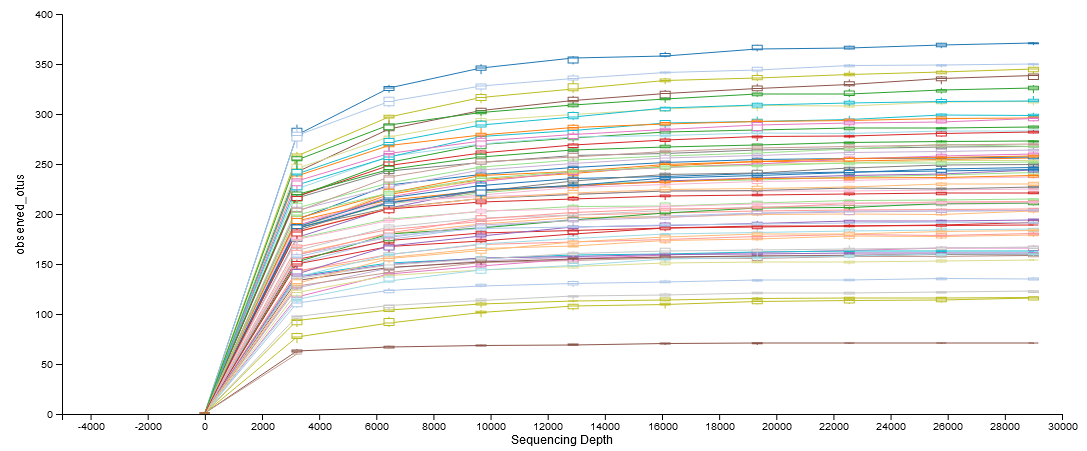

**Figure S3.** Rarefaction curve showing sample sequencing depth and that at the number used for rarefaction of samples (29,000 reads) diversity was consistent across samples.

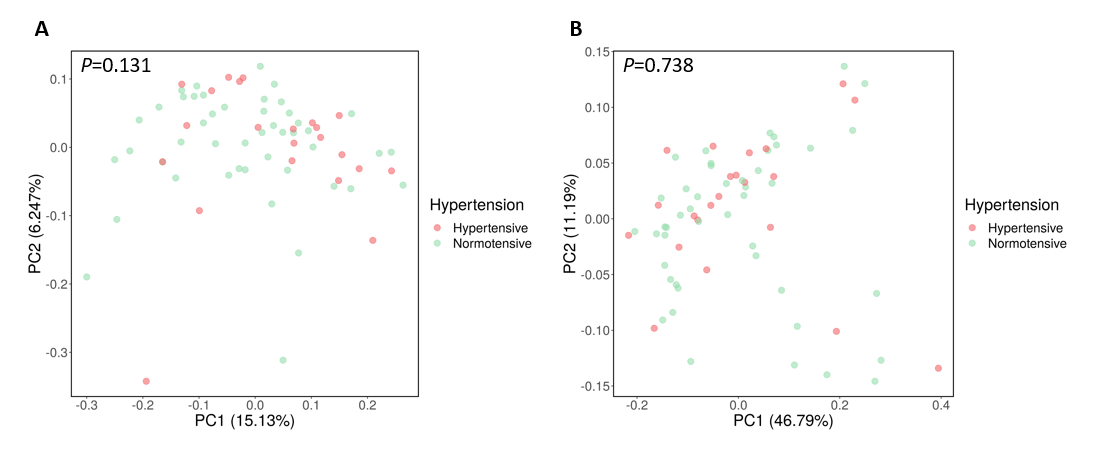

**Figure S4**. β diversity principal coordinate analysis plot showing no difference in the gut microbiome between normotensive (green) and hypertensive (red) participants. A, Unweighted (i.e., microbial diversity based on presence/absence) and B, weighted (i.e., microbial diversity based on abundance) UniFrac analyses.

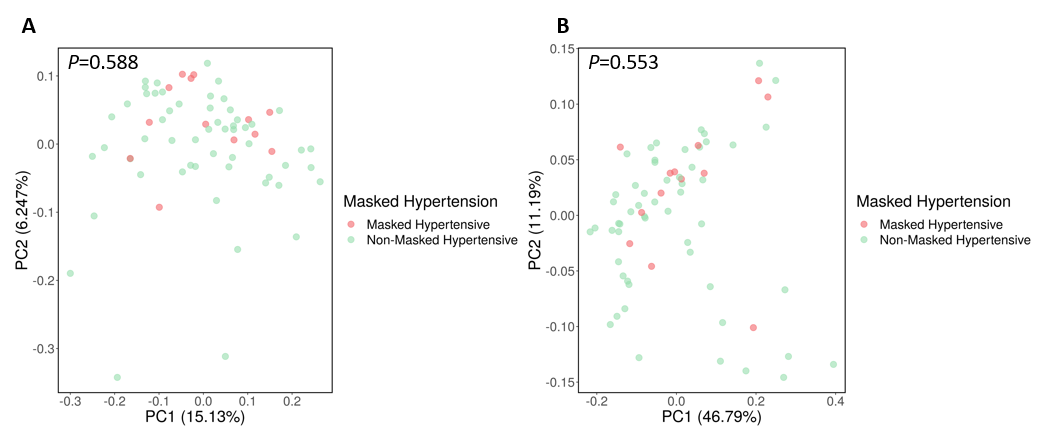

**Figure S5**. β diversity principal coordinate analysis plot showing no difference in the gut microbiome between normotensive (green) and masked hypertensive (red) participants. A, Unweighted (i.e., microbial diversity based on presence/absence) and B, weighted (i.e., microbial diversity based on abundance) UniFrac analyses.

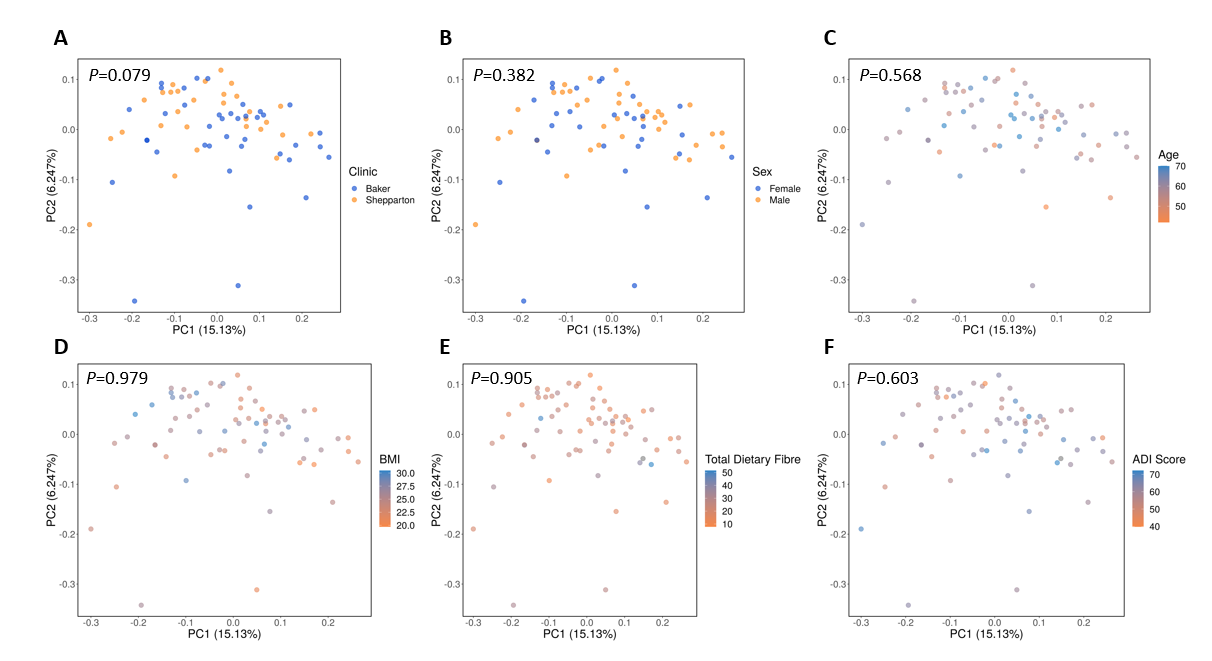

**Figure S6**. β diversity principal coordinate analysis plot showing no difference in the gut microbiome between A, clinics (Baker=metropolitan, Shepparton=regional), B, sexes, C, age, D, BMI, E, dietary fibre intake and F, Australia Dietary Index (ADI) score.

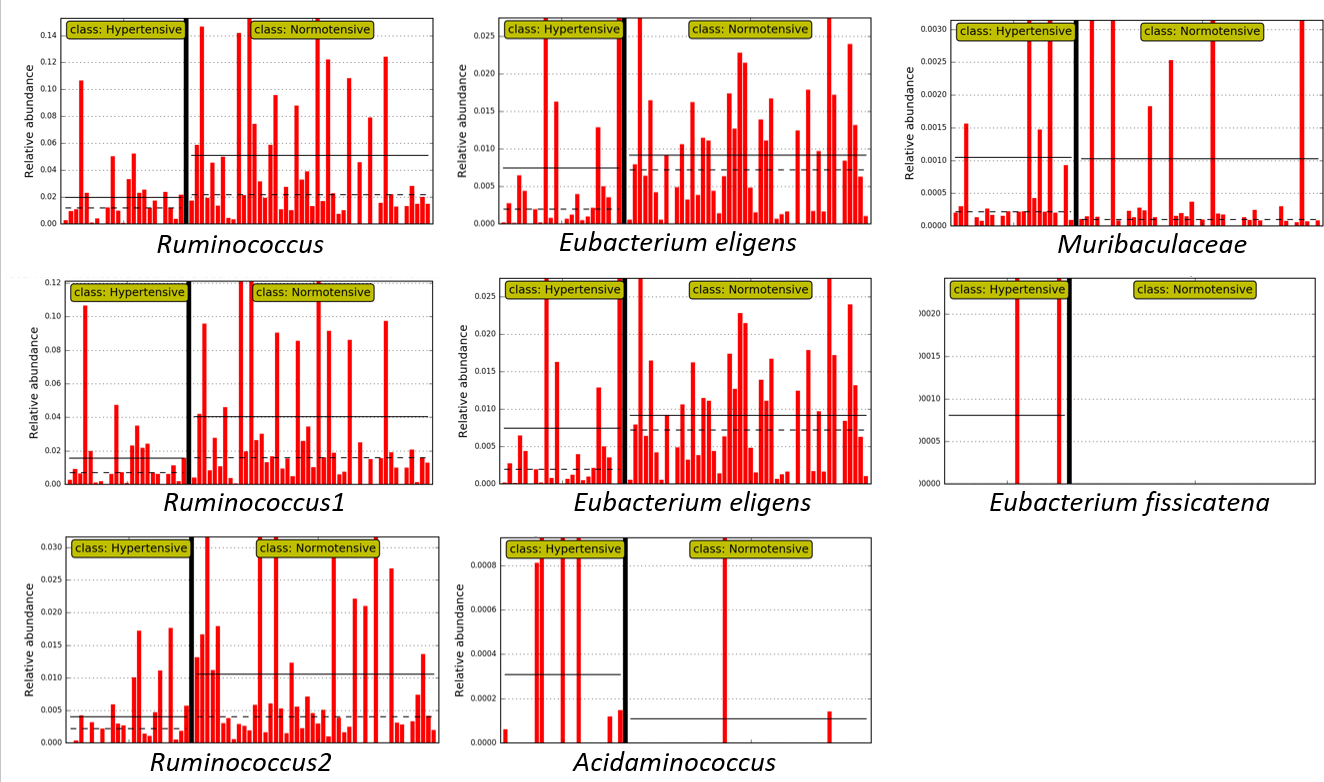

**Figure S7.** Levels of most prevalent taxa identified in LDA analysis between normotensives and essential hypertensives (reported in Figure 1). Straight horizontal line shows subclass means, dotted horizontal lines shows subclass medians.

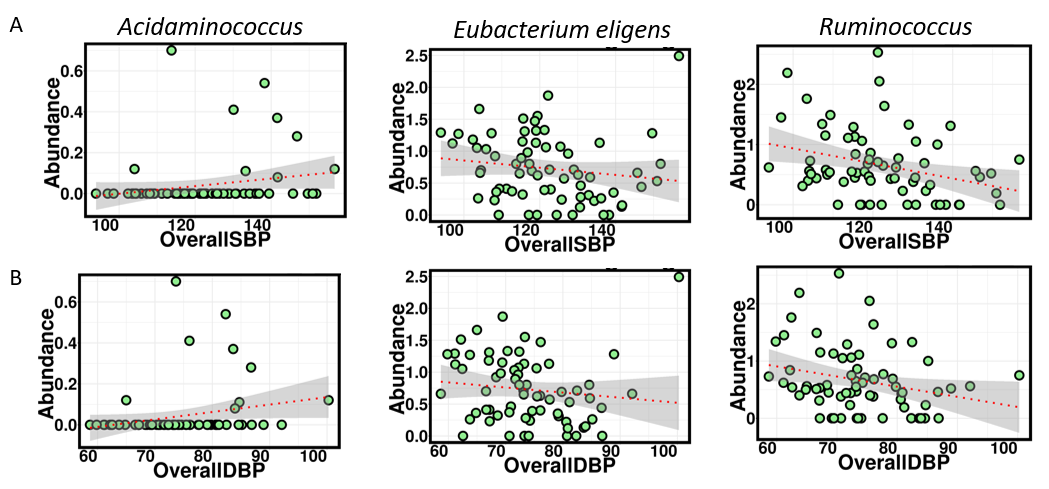
 **Figure S8**. Spearman correlation with 24-hour **A**, systolic and **B**, diastolic blood pressure (see Table S4 for r and P-values).

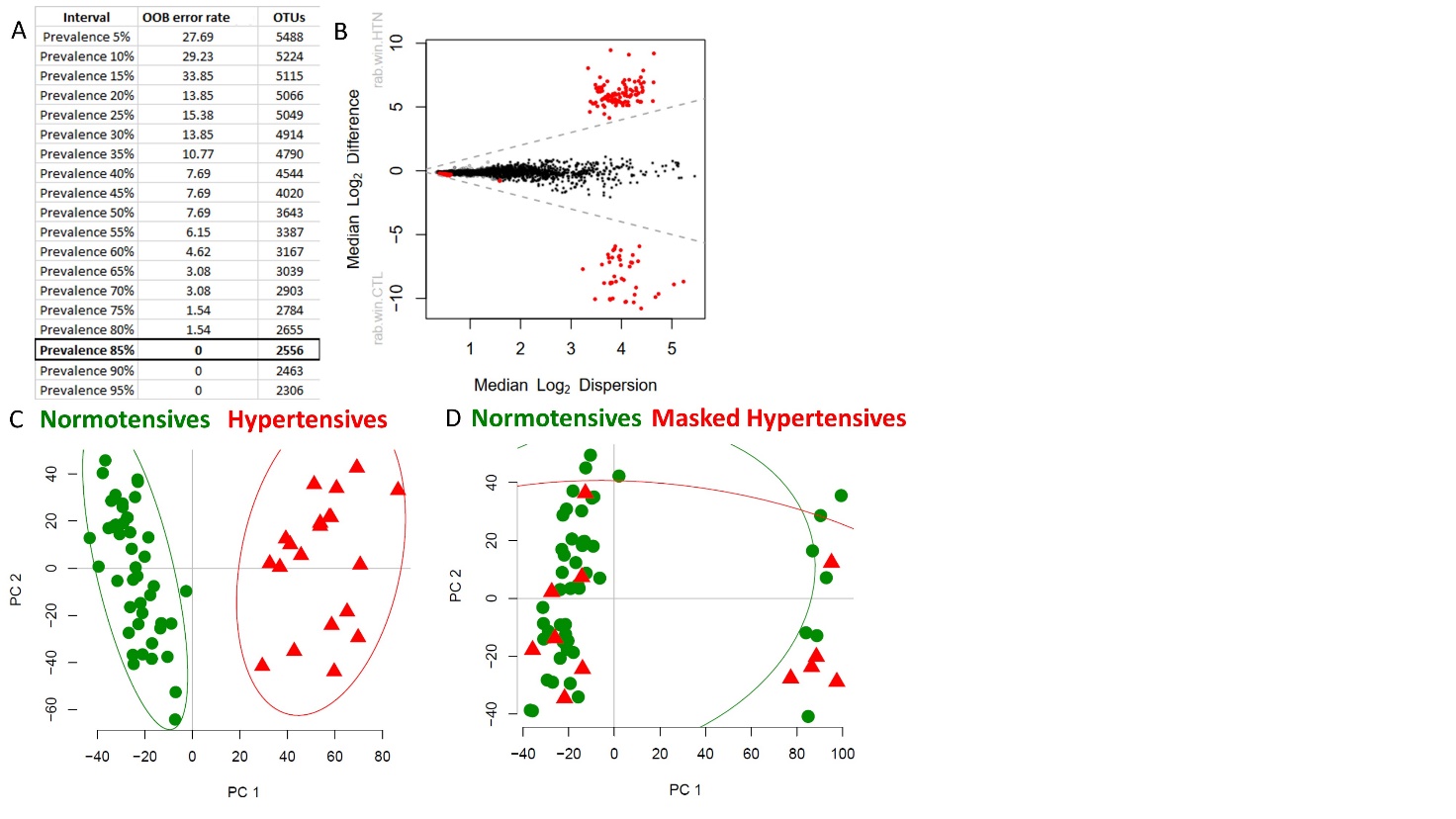

**Figure S9**. Using the DADA2/PIME/ALDEx2 machine-learning pipeline described in Methods (Piphillin server), the initial microbiome ASV dataset of 5,488 total raw data pathways was parsed into 5% intervals and ranked by out-of-bag (OOB) errors of signal-to-noise ratios giving minimized OBB error rate = 0 at the 85% prevalence level. This interval held 2,556 gene pathways. B. Mann-Whitney plot validating the 2,556 prevalent pathways (red points) of the 85% prevalence interval of (A). These pathways were subsequently used downstream for all the ensuing multivariate analyses comparing the groups. Points represent unique microbial gene functions that are differentially abundant at q<0.1 (red); abundant but non-differentially abundant (gray); rare and not differentially abundant (black). C. Principal component analysis (PCA) of prevalent microbial gene pathways using the data pool of (A) and (B), comparing essential hypertensive subjects (n=20) vs. normotensives (n=45). D. PCA comparing masked hypertensives (n=12) vs. normotensive and hypertensive subjects (n=57). All PCA points are shown within 95% confidence interval ellipses.

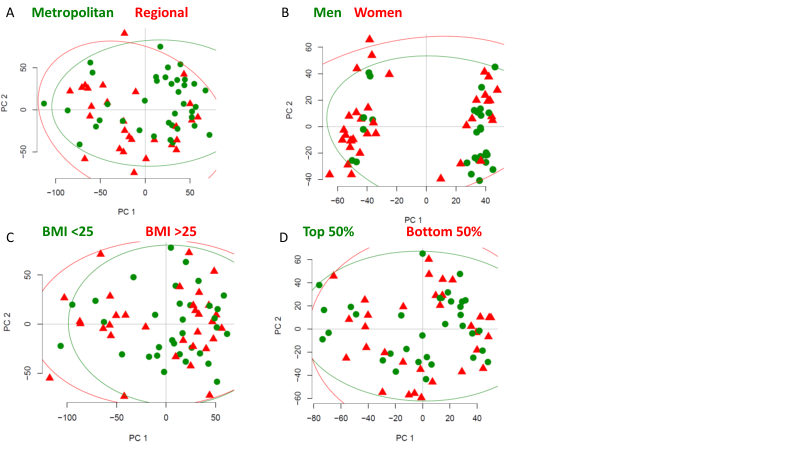

**Figure S10**. Pathway analyses plots showing no difference in the gut microbiome between **A**, clinics, **B,** sexes, **C,** BMI, **D,** Australia dietary score index. Data according to Piphillin server but validated in PICRUSt2 (Figure S10).

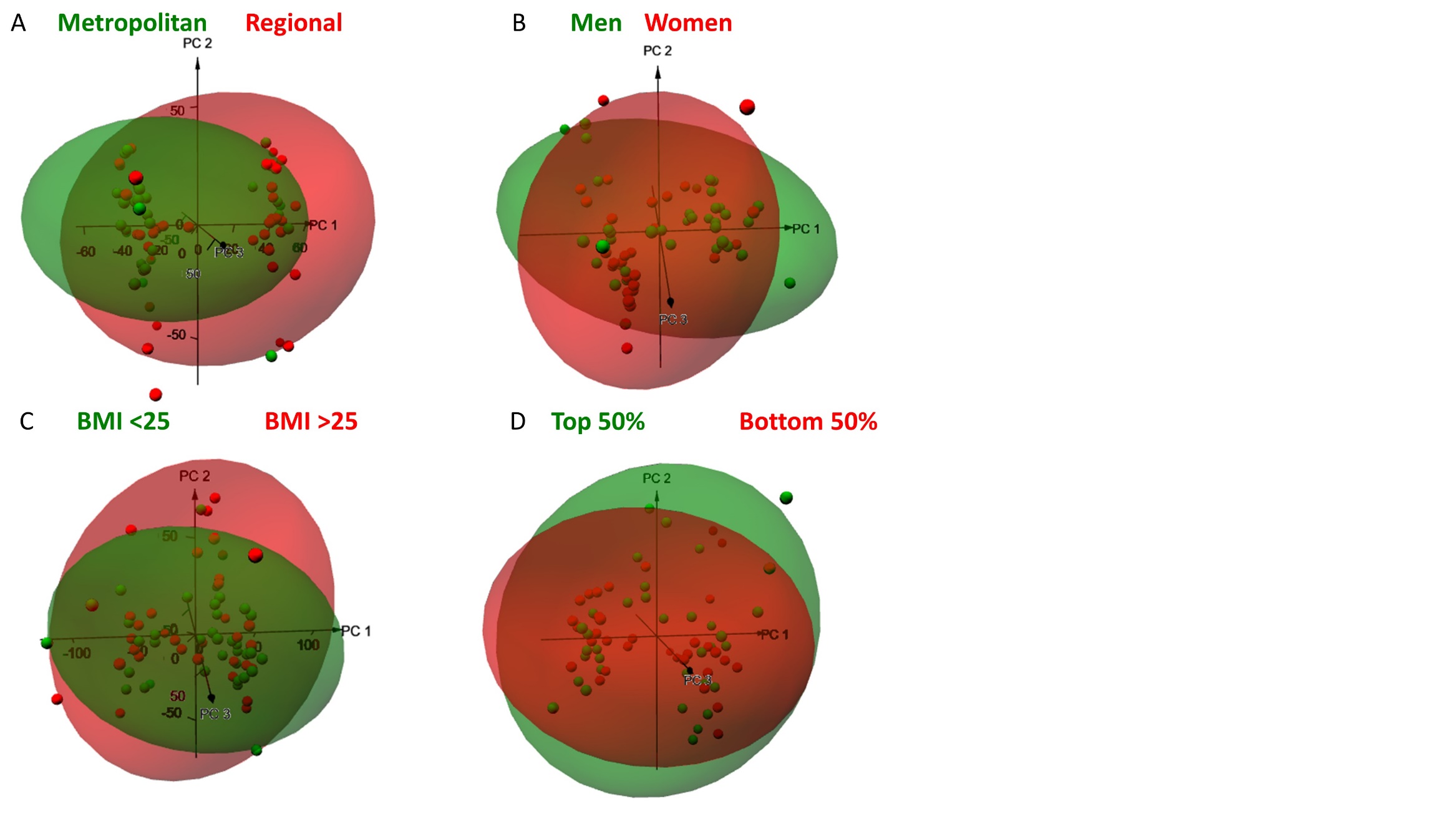

**Figure S11**. Pathway analyses plots showing no difference in the gut microbiome between **A**, clinics, **B,** sexes, **C,** BMI, **D,** Australia dietary score index. Data according to PICRUSt2 server but validated in Piphillin (Figure S9).

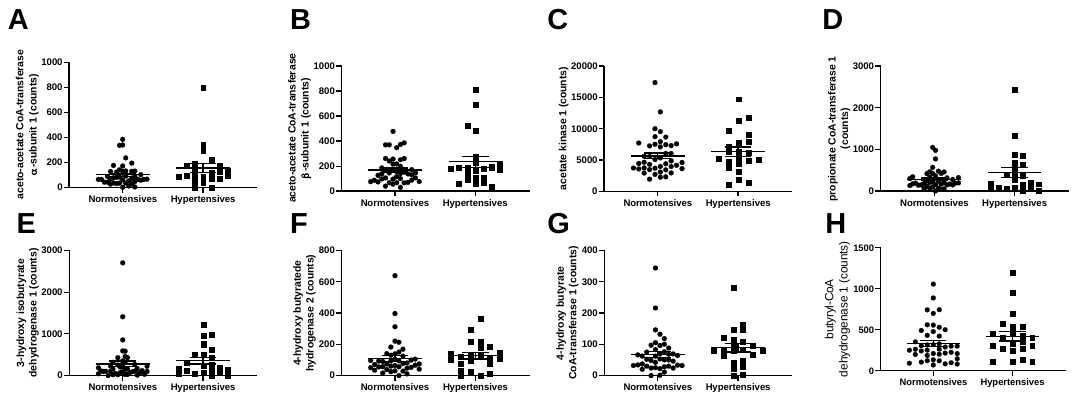
**Figure S12**. Pathways relevant to short-chain fatty acid production that were not differentially regulated between hypertensive and normotensive participants. Data according to Piphillin server but validated in PICRUSt2.
